# Trials underestimate the impact of preventive treatment for household contacts exposed to multidrug-resistant tuberculosis: a simulation study

**DOI:** 10.1101/2023.02.06.23285528

**Authors:** Parastu Kasaie, Jeff Pennington, Amita Gupta, David W. Dowdy, Emily A. Kendall

**Affiliations:** Department of Epidemiology, Johns Hopkins Bloomberg School of Public Health, Baltimore, Maryland; Division of Infectious Diseases, Johns Hopkins University School of Medicine, Baltimore, Maryland

**Keywords:** Tuberculosis, Multidrug-Resistant, Contact Tracing, Isoniazid, Computer Simulation, India

## Abstract

**Background:** Several clinical trials of tuberculosis preventive treatment (TPT) for household contacts of patients with multidrug-resistant tuberculosis (MDR-TB) are nearing completion. The potential benefits of TPT for MDR-TB contacts extend beyond the outcomes that clinical trials can measure.

**Methods:** We developed an agent-based, household-structured TB and MDR-TB transmission model, calibrated to an illustrative setting in India, the country accounting for 26% of global MDR-TB burden. We simulated household contact investigation for contacts of patients with MDR-TB, comparing an MDR-TPT regimen against alternatives of isoniazid preventive treatment, household contact investigation without TPT, or no household contact intervention. We simulated outcomes of a clinical trial and estimated the patient-level and population-level effects over a longer time horizon.

**Findings:** During two years of follow-up per recipient, a simulated 6-month MDR-TPT regimen with 70% efficacy against both DS- and MDR-TB infection could prevent 72% [Interquartile range (IQR): 45 – 100%] of incident MDR-TB among TPT recipients (number needed to treat (NNT) 73 [44 – 176] to prevent one MDR-TB case), compared to household contact investigation without TPT. This NNT decreased to 54 [30 – 183] when median follow-up was increased from two to 16 years, to 27 [11 – Inf] when downstream transmission effects were also considered, and to 12 [8 – 22] when these effects were compared to a scenario of no household contact intervention.

**Interpretation:** If forthcoming trial results demonstrate efficacy, the long-term population impact of MDR-TPT implementation could be much greater than suggested by trial outcomes alone.

**Funding:** NIH K01AI138853 and K08AI127908; Johns Hopkins Catalyst Award.

## Introduction

Household contacts of people diagnosed with tuberculosis (TB) benefit from TB screening and TB preventive treatment (TPT).^1,2^ Contact with rifampin- or multidrug-resistant TB (MDR-TB) carries additional risk, because delays in diagnosis and effective treatment of MDR-TB prolong exposure among contacts,^3^ and because the treatment options for contacts who develop MDR-TB are less effective and more toxic.^4^ Therefore, interventions to detect or prevent TB, including MDR-TB, in the contacts of MDR-TB patients are a high clinical and public health priority.

Despite the potential for benefit, efforts to intervene among contacts of people with MDR-TB have been limited by the lack of a TPT regimen with proven activity against MDR-TB. Now, however, WHO conditionally recommends preventive treatment for high-risk household contacts of people with MDR-TB,^5^ and at least three clinical trials will soon provide robust data on the efficacy of fluoroquinolones or delamanid as preventive treatment in this population.^6-8^

If these trials demonstrate efficacy of TPT in preventing MDR-TB among household contacts, the epidemiological impact of this TPT will extend beyond trial-based estimates of efficacy. TPT could both cure TB infections that would otherwise progress to TB disease after trial completion and prevent secondary transmission events that would otherwise cause TB in individuals who are not clinical trial participants. In addition, household contact investigation itself has important benefits, given that 4% or more of MDR-TB household contacts may have co-prevalent active TB^9^ and uptake of contact investigation remains low in many settings.^10^

In weighing risks and benefits in decisions about whether to scale up preventive treatment for MDR-TB contacts, it is important to capture this full spectrum of expected impact. We therefore developed an agent-based, household-structured transmission model of a TB epidemic, to better understand the relationship between trial-measured effects and anticipated population-level impact of TPT for MDR-TB household contacts.

## Methods

### Overview

We calibrated a TB transmission model to data from India, the country accounting for 26% of MDR TB cases in the world, and we used this model to simulate a preventive treatment intervention with efficacy against MDR-TB, implemented from 2023 to 2027. We compared the patient-level and population-level effects of this intervention to those of isoniazid preventive treatment, active TB screening alone, or no household contact intervention, over a time horizon of up to 18 years (through year 2040).

At an individual level, the natural history of TB is modeled as shown in **Figure 1A**. We model circulation of both drug-susceptible (DS-TB) and MDR-TB strains in the population. Individuals are born susceptible to TB infection. Upon infection (or reinfection) with DS-TB or MDR-TB, individuals move through an initial period of early latent infection (ELTB) with a gradually decreasing monthly risk of developing active TB disease via “primary progression”, followed by an indefinite period of late latent infection (LLTB) with a low but constant monthly risk of developing active TB via “reactivation”. Individuals with active TB may transmit infection to their contacts (both within and outside the household), experience increased mortality risk (not shown in **Figure 1**), and can recover from TB disease through treatment or spontaneous resolution; those who are recently recovered experience a risk of relapse for two years. Reflecting evidence that TB lesions in the same host may act independently,^11^ individuals may experience joint latent DS and MDR infections – but episodes of TB disease are modeled as either DS or MDR. Latently infected or recovered individuals (regardless of strain) experience partial protection against both DS-TB and MDR-TB reinfection. Individuals with active disease can undergo diagnosis and receive treatment, which may result in recovery, treatment failure (with continued TB mortality risk and partial infectiousness), or (for those initially with active DS-TB) acquisition of resistance and development of active MDR-TB at the end of treatment.

**Figure 1:**
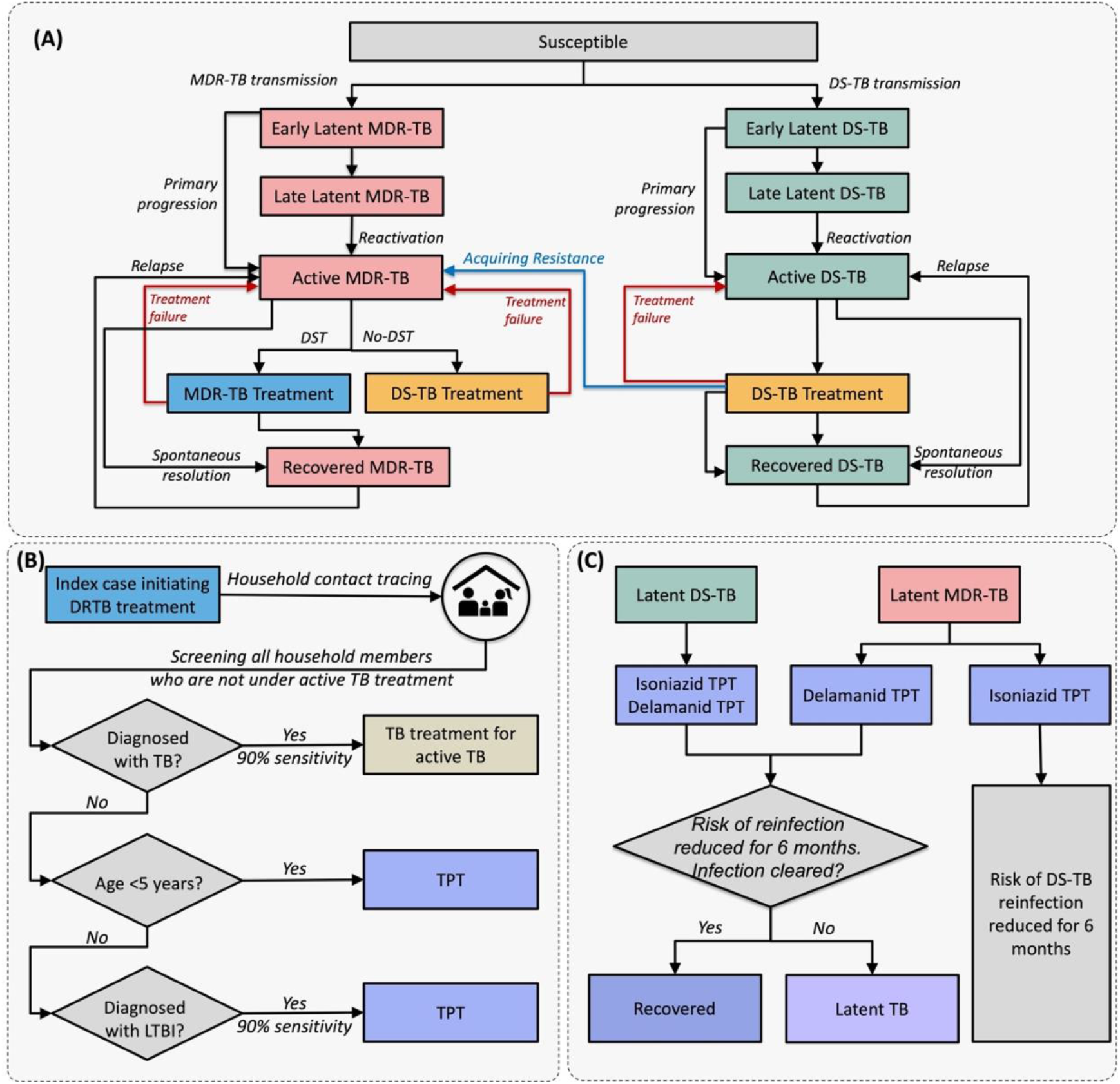
Schematic representation of TB natural history and modeled interventions. Panel A illustrates the natural history of drug-susceptible TB (DS-TB) and multidrug-resistant TB (MDR-TB); risk of mortality is included in the model but not shown here. Panel B illustrates the household contact investigation process. Household contacts diagnosed with active TB and those receiving TB treatment are ineligible for TPT. Panel C illustrates the modeled effects of isoniazid and delamanid TPT; prevention of relapse among recently recovered patients is also included but not shown here. Relapse risk is present only in the first two years after treatment of active TB and does not apply to those in the recovered state after preventive treatment.

Individuals with active TB have a probability of transmitting infection to the other members of their households (assigned based on household demographic data from India) and a lower probability of transmitting to their community contacts (randomly selected from the simulated population based on an age-structured contact matrix) at each time step. Details of model structure and parameterization are presented in **Appendix S1**.

### Setting and calibration

We simulate a growing population, with a size of approximately 800,000 people in 2022 and with demographics and TB burden based on those of India as a whole. To simulate historical development of an MDR-TB epidemic, the transmission model is first run without MDR-TB for a “burn-in” period of nearly 500 years, to reach a steady state reflective of the TB epidemic in 1970. MDR-TB acquisition is then allowed to occur after 1970 (reflecting the time at which rifampin became widely used), and effective MDR-TB detection and treatment are gradually introduced after 2007.^12^ Starting from prior distributions for each parameter (informed by literature where possible, and chosen to be minimally informative otherwise; **Table 1**), a sampling-importance-resampling process is used to calibrate the model to data from India on TB incidence and the temporal trend in incidence,^12^ TB mortality,^12^ the proportions of notified new and previously-treated TB patients that have drug-resistant disease,^12^ the prevalence of latent TB infection (LTBI),^13^ and the prevalence of active TB^14^, as well as to an estimate from high-burden settings of the proportion of active TB that results from household-based transmission^15^ (**Table S3**).

**Table 1:**
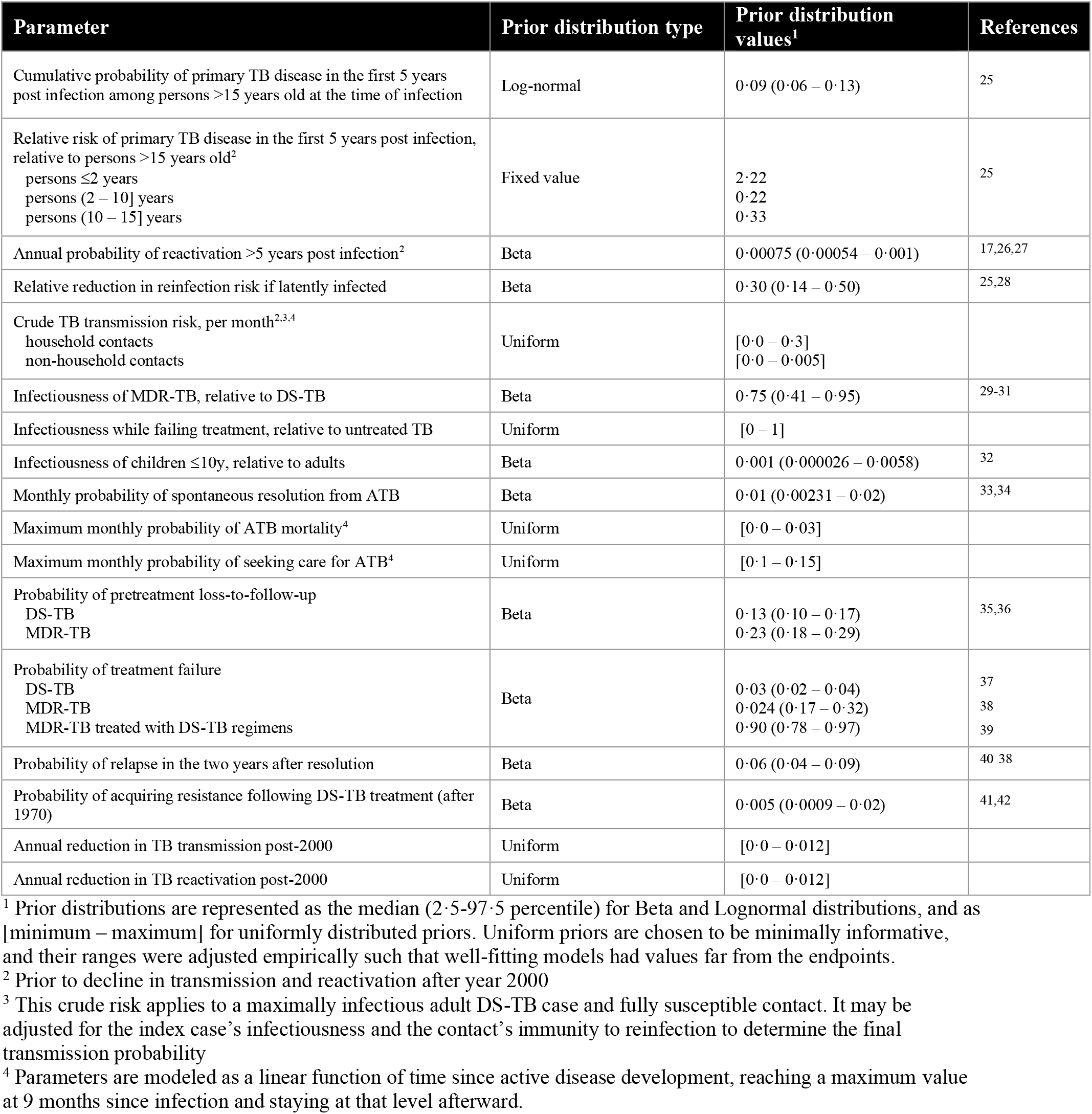
List of model parameters and prior distributions.

| Parameter | Prior distribution type | Prior distribution values<sup>1</sup> | References |
| --- | --- | --- | --- |
| Cumulative probability of primary TB disease in the first 5 years post infection among persons >15 years old at the time of infection | Log-normal | 0.09 (0.06 – 0.13) | 25 |
| Relative risk of primary TB disease in the first 5 years post infection, relative to persons >15 years old<sup>2</sup><br>persons ≤2 years<br>persons (2 – 10] years<br>persons (10 – 15] years | Fixed value | 2.22<br>0.22<br>0.33 | 25 |
| Annual probability of reactivation >5 years post infection<sup>2</sup> | Beta | 0.00075 (0.00054 – 0.001) | 17,26,27 |
| Relative reduction in reinfection risk if latently infected | Beta | 0.30 (0.14 – 0.50) | 25,28 |
| Crude TB transmission risk, per month<sup>2,3,4</sup><br>household contacts<br>non-household contacts | Uniform | [0.0 – 0.3]<br>[0.0 – 0.005] |  |
| Infectiousness of MDR-TB, relative to DS-TB | Beta | 0.75 (0.41 – 0.95) | 29-31 |
| Infectiousness while failing treatment, relative to untreated TB | Uniform | [0 – 1] |  |
| Infectiousness of children ≤10y, relative to adults | Beta | 0.001 (0.000026 – 0.0058) | 32 |
| Monthly probability of spontaneous resolution from ATB | Beta | 0.01 (0.00231 – 0.02) | 33,34 |
| Maximum monthly probability of ATB mortality<sup>4</sup> | Uniform | [0.0 – 0.03] |  |
| Maximum monthly probability of seeking care for ATB<sup>4</sup> | Uniform | [0.1 – 0.15] |  |
| Probability of pretreatment loss-to-follow-up<br>DS-TB<br>MDR-TB | Beta | 0.13 (0.10 – 0.17)<br>0.23 (0.18 – 0.29) | 35,36 |
| Probability of treatment failure<br>DS-TB<br>MDR-TB<br>MDR-TB treated with DS-TB regimens | Beta | 0.03 (0.02 – 0.04)<br>0.024 (0.17 – 0.32)<br>0.90 (0.78 – 0.97) | 37<br>38<br>39 |
| Probability of relapse in the two years after resolution | Beta | 0.06 (0.04 – 0.09) | 40 38 |
| Probability of acquiring resistance following DS-TB treatment (after 1970) | Beta | 0.005 (0.0009 – 0.02) | 41,42 |
| Annual reduction in TB transmission post-2000 | Uniform | [0.0 – 0.012] |  |
| Annual reduction in TB reactivation post-2000 | Uniform | [0.0 – 0.012] |  |

<sup>1</sup> Prior distributions are represented as the median (2.5-97.5 percentile) for Beta and Lognormal distributions, and as [minimum – maximum] for uniformly distributed priors. Uniform priors are chosen to be minimally informative, and their ranges were adjusted empirically such that well-fitting models had values far from the endpoints.
<sup>2</sup> Prior to decline in transmission and reactivation after year 2000
<sup>3</sup> This crude risk applies to a maximally infectious adult DS-TB case and fully susceptible contact. It may be adjusted for the index case's infectiousness and the contact's immunity to reinfection to determine the final transmission probability
<sup>4</sup> Parameters are modeled as a linear function of time since active disease development, reaching a maximum value at 9 months since infection and staying at that level afterward.

### Experimental scenarios

We simulate a household contact tracing (HHCT) intervention in the households of patients with active MDR-TB (**Figure 1, Panel B**). When a simulated patient initiates treatment for MDR-TB, all household members not already on TB treatment are screened for active TB (with 90% sensitivity), and those with active TB are treated with a DS- or MDR-TB regimen as appropriate. Contacts who are under five years old, or older than five with evidence of LTBI (assuming 90% sensitivity of testing for LTBI), receive TPT.

To evaluate different TPT strategies and also reflect the ongoing PHOENIx trial,^8^ we compare four different scenarios:

1. a 6-month TPT regimen with activity only against DS-TB infections (“isoniazid”);
2. a 6-month TPT regimen with activity against both DS and MDR-TB infections (“delamanid”, although results should apply to other TPT regimens with demonstrated activity against MDR-TB);
3. a comparator in which household contacts are screened for active TB but not given TPT (“placebo”); and
4. a no-intervention comparator without HHCT (“no screening”).

The intervention is modeled as beginning in year 2023 and continuing for 5 years; during that period, the modeled intervention is delivered to 70% of all intervention-naÏve households of patients who initiate treatment for MDR-TB. TPT recipients and the broader population are followed to 2040 to project longer-term impacts on epidemiological outcomes.

### Effects of preventive treatment

TPT is modeled as having three effects on TB risk (**Figure 1C**). Due to imperfect efficacy and adherence, each effect is modeled as occurring with 70% probability (varied in sensitivity analysis) for TB strains against which the drug has activity. First, TPT prevents progression from latent infection to active disease; this is modeled as moving a proportion of recipients with latent TB (regardless of time since infection) to the recovered state, without risk of relapse. This effect thus persists after TPT, so long as reinfection does not occur. Second, among individuals who have recently recovered from active TB, TPT prevents relapse events that would otherwise occur. Third, for individuals who are exposed to *M. tuberculosis* while taking TPT, TPT prevents infection/reinfection; this effect ends with TPT completion. For each of these three effects, we assume that isoniazid TPT affects only DS strains, that delamanid has the same effects on both DS and MDR strains, and that TPT has no effect against active TB disease (for example, if missed during screening).

### Sensitivity analysis

For each sampled parameter in **Table 1**, we used partial rank correlation coefficients (PRCCs) to describe the association between that parameter and the projected reduction in cumulative, population-wide MDR-TB incidence produced by HHCT plus delamanid TPT by 2040, relative to no household intervention. The parameters most strongly correlated with this primary outcome (i.e., absolute value of PRCC>0·15) were selected for one-way sensitivity analysis, in which we compared outcomes between the 20% of calibrated simulations with the highest values for that parameter versus the 20% with the lowest values for that parameter. We also assessed the dependence of estimated TPT impact on assumptions regarding the intervention, including the efficacy values assigned to each of the three modeled mechanisms of TPT effect.

## RESULTS

The model recapitulated most epidemiological targets in 2019 (**Figure 2**). In the year 2022 (pre-intervention baseline), a median of 0·28% [Interquartile Range (IQR): 0·27 – 0·29%] of the modeled population had prevalent active TB (i.e., 280 per 100,000 population), and 24% [IQR 22 – 32%] of incident TB arose from transmission within households (**Figure 2**). Of all incident TB, 3·2% [IQR 2·7 – 3·6%] was drug-resistant. In the absence of additional intervention, TB incidence was projected to fall by 5·1% [IQR 3·9 – 5·3%] per year between 2022 and 2040 (**Table S5**).

**Figure 2:**
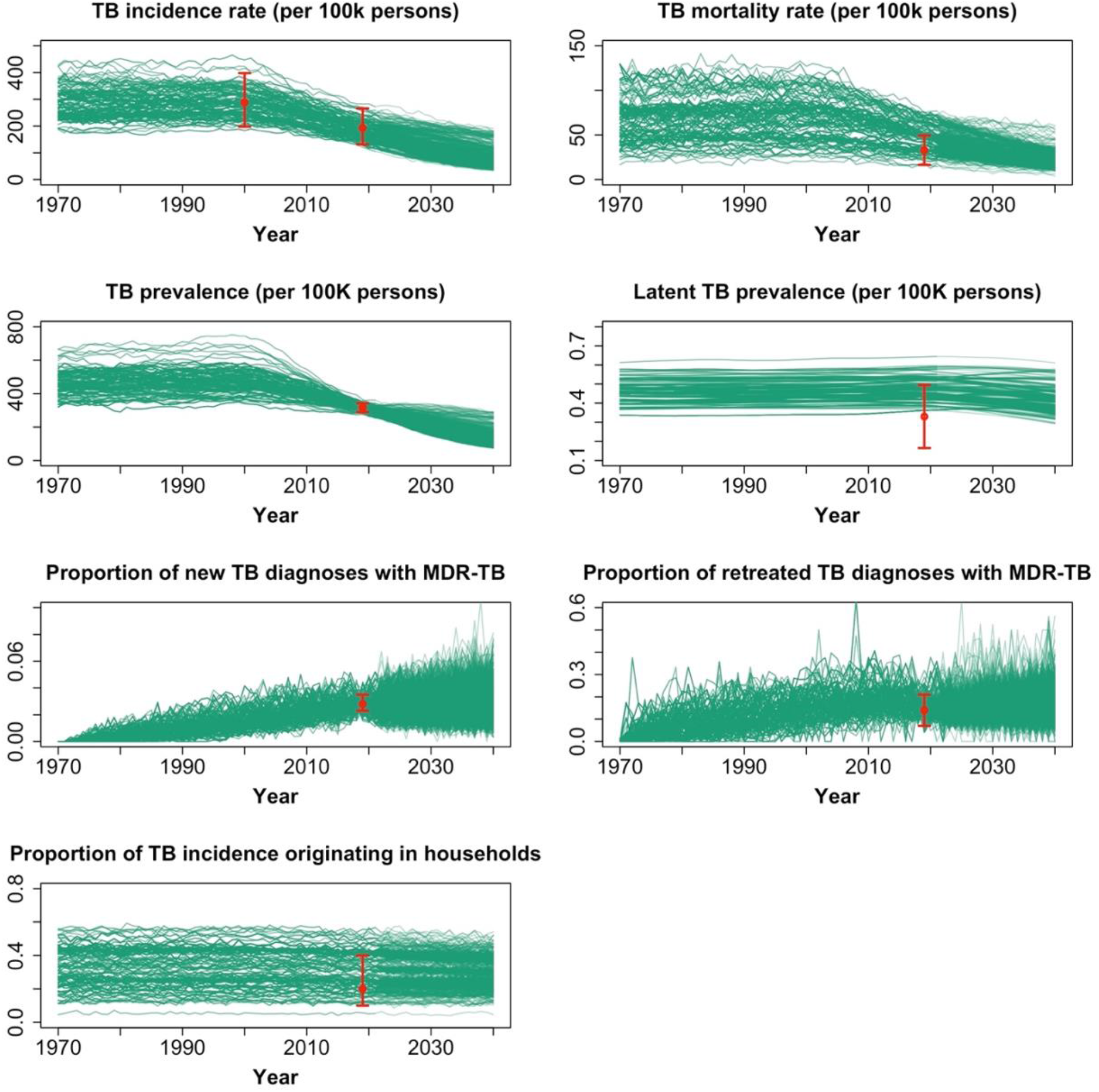
Calibration targets and simulated epidemic trajectories. The green curves represent the values for each parameter listed, among the 1000 best-fitting simulations from 1970 through 2040 and assuming no interventions for MDR-TB household contacts (i.e., no-intervention scenario). Red dots and error bars represent the median and inner 0·95 quantile range, respectively, of each calibration target (i.e., data to which the model was fit).

Within the households of all patients diagnosed with TB in 2022, the median estimated prevalence of LTBI was 69% [IQR 68 – 77%], and the co-prevalence of active TB was 3·6% [IQR 3·4 – 4·9%]. Considering only those households where the index case had MDR-TB, the estimated prevalence of LTBI was 74% [IQR 73 – 78%], and the co-prevalence of active TB was 3·9% [IQR 3·5 – 4·8%]. Among latently-infected individuals in these MDR-TB contact households, 62% [IQR 60% – 67%] had an MDR infection, and most of their latent MDR infections were still in the early latent stage with high progression risk (89% [IQR 86-90%], in contrast to 27% [IQR 25 – 29%] of DS infections in the same households). For the active TB that was present at the time of HHCT in these in these same MDR-TB contact households, an even greater proportion (81% [IQR 71% – 98%]) was MDR.

### Simulated outcomes of an MDR-TB preventive treatment trial

Between 2023 and 2027, in simulated populations of approximately 800,000 people each, a median of 169 [IQR 134– 178] people developed incident MDR-TB, and 123 [IQR 112 – 127] people were diagnosed with MDR-TB. Delivering contact investigation to the households of 70% of these 123 patients required screening a median of 314 [IQR 255 – 327] household contacts. Among these contacts, 10 [IQR 8 –11] were diagnosed with active TB, of whom 80% [IQR 78 – 83%] had MDR-TB. An additional 211 [IQR 175 – 225] contacts were identified as eligible for preventive treatment: 34 [IQR 29 – 36] contacts under age 5, and 176 [IQR 146 – 189] older contacts with evidence of TB infection (**Table S6**).

In the “placebo” scenario, the projected 2-year cumulative incidence of DS-TB among TPT-eligible MDR-TB contacts was 0·4% [IQR 0 – 0·8%], and the 2-year cumulative incidence of MDR-TB was 2% [IQR 1 – 3%] (**Table 2**). As illustrated for one simulated trial in **Figure 3**, isoniazid TPT reduced the two-year cumulative incidence of DS-TB to 0 [0 – 0·4%], with negligible impact on MDR-TB incidence, while delamanid TPT additionally reduced the two-year incidence of MDR-TB to 0·6% [IQR 0 – 1·1%], a 71% [IQR 45 – 100%] reduction. From the perspective of a two-year clinical trial, therefore, the estimated number needed to treat (NNT) with delamanid TPT to prevent one TB case relative to placebo was 60 [IQR 37 – 130], and the NNT to prevent one MDR-TB case was 73 [IQR 44 – 176].

**Table 2:** Simulated impact of delamanid TPT for household contacts of people diagnosed with MDR-TB, from a clinical trial perspective and increasingly comprehensive perspectives.

| **Perspective** | **Population under consideration<sup>1</sup>** | **Comparator scenario** | **Median duration of follow up** | **MDR-TB cases in comparator scenario (per TPT recipient)** | **MDR-TB cases prevented by delamanid TPT** | **% of MDR-TB cases prevented among the population under consideration** | **NNT to prevent one MDR-TB case** |
| --- | --- | --- | --- | --- | --- | --- | --- |
| **Clinical trial** | TPT recipients | Placebo | 2 years | 0.02<br>[0.01 – 0.03] | 0.01<br>[0.006 – 0.02] | 72%<br>[45 – 100%] | **73 [44 – 176]** |
| **Extended clinical trial follow up** | TPT recipients | Placebo | 16 years | 0.04<br>[0.03 – 0.05] | 0.02<br>[0.01 – 0.03] | 51%<br>[20 – 72%] | **54 [30 – 183]** |
| **Including downstream transmission effects** | Full population | Placebo | 16 years | 2.2 [1.9 - 2.4] | 0.04 [-0.01 - 0.09] | 1.6%<br>[-0.6 – 3.8%] | **27 [11 – Inf<sup>2</sup>]** |
| **Including effects of TB screening for contacts** | Full population | No household contact screening | 16 years | 2.3 [1.9 - 2.4] | 0.08 [0.04 - 0.12] | 3.6%<br>[2 – 5.2%] | **12 [8 – 22]** |
| **Perspective** | **Population under consideration<sup>1</sup>** | **Comparator scenario** | **Median duration of follow up** | **TB cases in comparator scenario (per TPT recipient)** | **TB cases prevented by delamanid TPT** | **% of TB cases prevented among the population under consideration** | **NNT to prevent one TB case** |
| **Clinical trial** | TPT recipients | Placebo | 2 years | 0.02<br>[0.02 – 0.04] | 0.02<br>[0.01 – 0.03] | 71%<br>[46 – 88%] | **60 [37 – 130]** |
| **Extended clinical trial follow up** | TPT recipients | Placebo | 16 years | 0.06<br>[0.04 – 0.08] | 0.03<br>[0.01 – 0.04] | 45%<br>[18 – 64%] | **40 [23 – 124]** |
| **Including downstream transmission effects** | Full population | Placebo | 16 years | 70.2 [67.7 - 87.6] | 0.05 [-1.4 – 1.6] | 0.08%<br>[-1.8 – 2%] | **18 [1 – Inf<sup>2</sup>]** |
| **Including effects of TB screening for contacts** | Full population | No household contact screening | 16 years | 70.4 [67.8 - 87.2] | 0.13 [-0.15 - 0.39] | 0.19%<br>[-0.19 – 0.52%] | **7 [3 – Inf<sup>2</sup>]** |

<sup>1</sup> TPT recipients are followed to incident TB or death, while the full population perspective counts all incident TB events including recurrences.
<sup>2</sup> A 75th percentile of “Inf” indicates that no TB is prevented in the overall population in >25% of simulations, due to stochastic effects in the large TB epidemic relative to the small number of households participating in the MDR-TB contact tracing intervention.
TB: tuberculosis; MDR: multidrug-resistant; TPT: TB preventive treatment; NNT: number needed to treat with TPT

**Figure 3:**
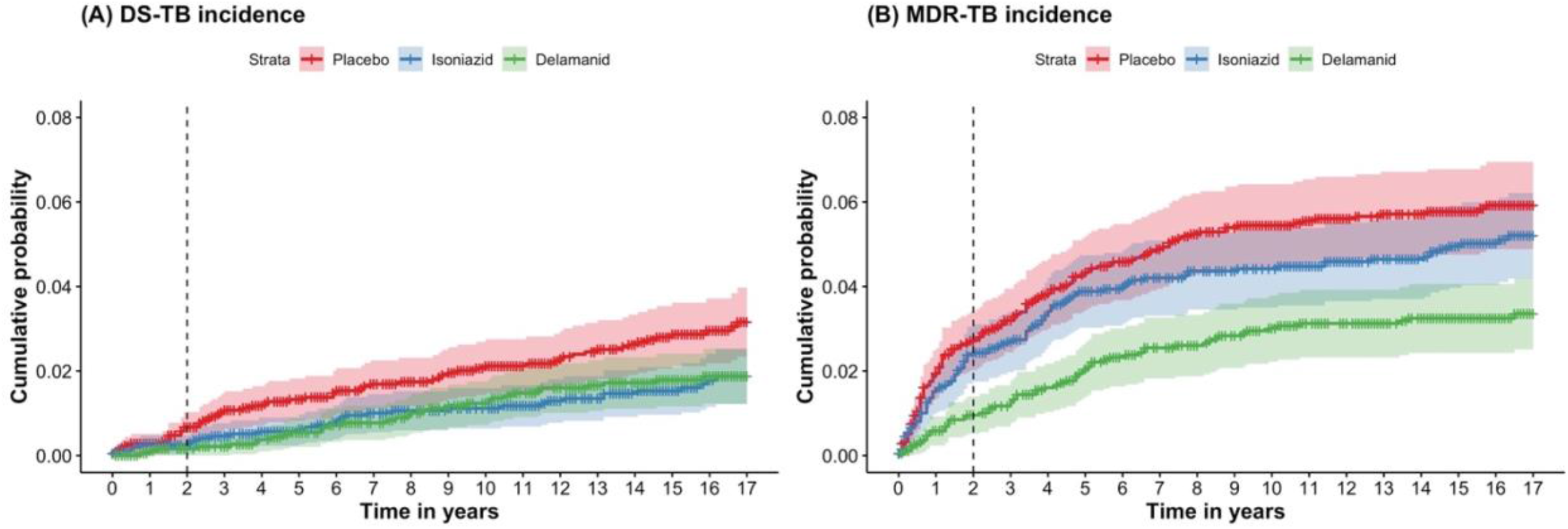
Projected TB incidence among TPT recipients in a simulated trial. For each scenario, 2000 individual TB preventive treatment (TPT) recipients per arm (representing one realization of a clinical trial) are randomly selected from across all simulations, their TPT start times are aligned, and they are followed until they either develop TB with the specified strain (drug-susceptible [DS] in panel A; multidrug-resistant [MDR] in panel B) or are censored due to death, development of active TB with the other resistance profile, or end of follow-up at year 2040. Colored ribbons represent two-sided 95% uncertainty ranges (R package survfit).

### Impact of MDR-TB preventive treatment from broader perspectives

When follow-up of TPT-eligible contacts was extended to 2040 (median 16·2 years of follow up), TPT-eligible MDR-TB contacts experienced a cumulative DS-TB incidence of 1·7% [IQR 0·9 – 2·6%] and cumulative MDR-TB incidence of 4% [IQR 3 – 5%] in the placebo scenario (**Table 2**; illustrated for one simulated trial in **Figure 3**). The majority of incident TB among these contacts continued to be MDR, although the proportion that was MDR decreased from 86% [IQR 71 – 100%] in the first two years to 69% [IQR 56 – 82%] in the remainder of this follow-up period.

Over this longer time horizon, recipients of delamanid TPT experienced a 42% [IQR-13 – 75%] reduction in the cumulative incidence of DS-TB (also experienced by isoniazid recipients) and a 51% [IQR 20 – 72%] reduction in the incidence of MDR-TB. Thus, among TPT-eligible contacts, the 16-year NNT to prevent one TB case with delamanid was 40 [IQR 23 – 124] and the NNT to prevent one MDR-TB case was 54 [IQR 30 – 183] (**Table S7**).

When also considering potential reductions in population-wide transmission, delivering delamanid TPT to 70% of eligible MDR-TB household contacts from 2023 through 2027 prevented 1·6% [IQR −0·6 – 3·8%] of all incident MDR-TB cases in the population between 2023 and 2040, relative to the placebo scenario (**Table 2** and **Figure 4**). This incorporation of transmission-related effects decreased the NNT to prevent one MDR-TB case to 27 [IQR 11 – infinite].

**Figure 4:**
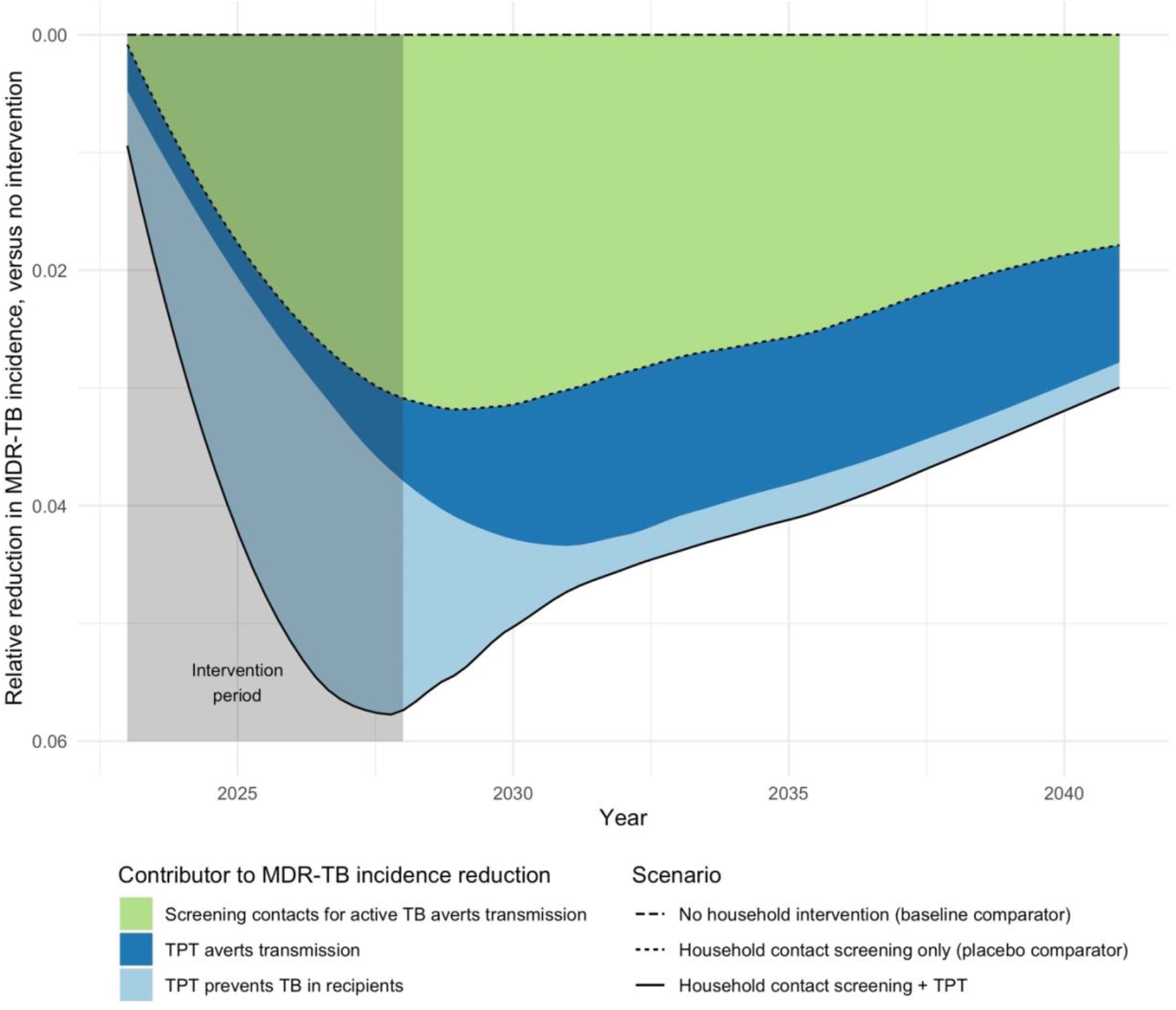
Projected reductions in MDR-TB incidence via household contact intervention. The Y-axis represents the reduction in MDR-TB incidence relative to the no-intervention baseline in the same year (a TPT-recipient-weighted average across simulations, smoothed using locally estimated scatterplot smoothing with a span of 0·5 years). The light blue region shows the effects that could be observed in a placebo-controlled clinical trial with sufficiently long follow up, and the dark blue shows additional effects of TPT in preventing transmission in the broader population; together, these blue regions represent the difference between the TPT scenario (solid line) and placebo (dotted line). The green region – the difference between the placebo scenario (dotted line) and no household intervention (dashed line) – represents the additional effect of active TB screening in the household contacts of MDR-TB patients.

Finally, when compared to a scenario of no household intervention (rather than screening for active TB without providing TPT, as in the placebo scenario), the combination of screening and delamanid TPT for MDR-TB household contacts from 2023 through 2027 averted a median of 3·6% [IQR 2 – 5·2%] of all incident MDR-TB cases in the population from 2023 to 2040 (**Figure 4**) – with a corresponding NNT of 12 [IQR 8 – 22] to prevent one MDR-TB case (**Table 2 & Table S8**).

### Sensitivity analysis

The epidemiologic parameters most strongly correlated with the primary outcome (relative reduction in MDR-TB incidence, comparing delamanid to no intervention) included the household TB transmission risk, the reduction in reinfection risk if latently infected, the TB mortality rate, the infectiousness of MDR-TB relative to DS-TB, and the primary progression rate. Of these parameters, only a high transmission risk among household contacts resulted in a >20% change in the projected impact of contact investigation plus delamanid TPT when varied across pre-specified ranges. Among intervention-related parameters, the impact of delamanid TPT among recipients was most sensitive to variation in the efficacy of TPT in clearing latent infections and the sensitivity of screening for active TB. Projected population-level effects were most sensitive to the coverage of household contact tracing and the sensitivity of LTBI screening (see **Appendix S3**).

## Discussion

This agent-based, household-structured model of MDR-TB transmission illustrates that the observed effect of TPT for MDR-TB household contacts in a clinical trial may substantially underestimate the long-term, population-wide impact of such an intervention. Specifically, we estimate that household contact investigation to screen for active TB and provide TPT with 70% efficacy against both DS- and MDR-TB infection could prevent one case of TB for every 12 [IQR 8 – 22] TPT recipients over an 18-year horizon. Less than 20% of this impact would be directly observable among TPT recipients themselves within 2 years of treatment. The remainder of this impact, which would not be measurable in a clinical trial with two-year follow-up, reflects the preventive effects among recipients over the longer term, the downstream effects of preventing MDR-TB transmission from those recipients to the broader population, and the benefits of screening and treating MDR-TB contacts for TB disease during implementation of household contact investigation.

Of these multiple contributions to the impact of MDR-TB household contact tracing and TPT, the largest contributor (accounting for 56% of the total estimated effect) was screening and treating contacts for active disease. In other words, much of the potential impact of MDR-TPT reflects the fact that having an effective TPT regimen (and newer effective regimens for MDR-TB treatment) might facilitate broader scale-up of contact investigation in the first place. Nevertheless, even in settings where TB screening for MDR-TB contacts is already widely implemented, the incremental benefit of adding TPT for this population could be both substantial (one MDR-TB case prevented per 27 TPT recipients) and considerably underestimated (by more than a factor of two) in clinical trials.

Despite its large benefit on a per-recipient basis, contact investigation with preventive treatment for MDR-TB is expected to have a relatively small impact on the overall incidence of MDR-TB in the population. This limited impact largely reflects the relatively small percentage of TB transmission that occurs within households,^16^ together with gaps in the cascade of care (e.g., imperfect sensitivity of LTBI tests, imperfect uptake and efficacy of TPT), the short five-year duration of the modeled intervention, and the large proportion of secondary household cases that already have active TB at the time of contact investigation.^2,17^ Nevertheless, for each household contact who receives effective MDR-TPT, the expected clinical benefits – for the TPT recipient and their future contacts – are substantial.

Among models of TB transmission, ours is distinctive in its representation of individual agents and dynamic household structures, together with co-circulation of DS and MDR strains. Including these details, while adding substantially to model complexity, is essential to accurately simulate household contacts’ time-varying risks of incident DS- and MDR-TB and the resulting effects of household-directed TB prevention. Our model accurately replicates observational data from household contact studies that were not used for calibration. For example, the simulated co-prevalence of active disease in households at the time of contact investigation (3·6% [IQR 3·4 – 4·9%]) is consistent with estimates ranging from 2·87% to 3·29% in previous systematic reviews^17-19^; the prevalence of TB infection among MDR-TB contacts (69% [IQR 68 – 77%]) is consistent with the 72% prevalence observed a multi-country feasibility study for an ongoing TPT trial ^2^; and the simulated strain concordance of co-prevalent TB cases found during MDR-TB contact investigation (81% [IQR 70 – 98%] and among MDR-TB contacts developing incident TB in the two years following contact investigation (80% [IQR 78 – 83%]) is consistent with the 82·6% (72·3-90·9%) isoniazid and rifampin concordance estimated in a systematic review of secondary cases in the households of patients with drug-resistant TB.^20^ Compared to related modeling analyses, our estimate that delamanid TPT prevents ∼51% of incident MDR-TB among all TPT recipients is consistent with a previously published estimate of ∼55% incidence reduction among pediatric contacts.^21^. At a population level, our median estimated NNT of 18 MDR-TB contacts receiving preventive treatment to prevent one TB case falls is comparable to the estimated NNT with conventional preventive treatment for DS-TB among people with HIV in South Africa (NNT=18) ^22^ and lower than a comparable estimate (NNT=64) for household contacts of DS-TB patients in Southeast Asia.^23^

As with any modeling study, this analysis has important limitations. We did not explicitly model HIV, making it difficult to generalize our results to high-HIV-burden settings or HIV-focused TPT strategies. We considered only DS and MDR strains; as such, we do not represent any effects of isoniazid monoresistance during MDR-TB emergence, nor any potential effects of TPT on MDR-TB susceptibility profiles over time. Our household-structured simulation represents household-specific and community-wide exposure risks, but it does not capture other risks that members of the same household may share, such as common non-household contacts or predisposing factors such as malnutrition. Our Bayesian calibration approach captures parameter uncertainty that is often omitted from agent-based simulation models, but it assumes a single set of parameter values within each simulation and thus does not capture inter-individual heterogeneity or dynamic changes in parameter values over time. For example, our model’s high estimate of the prevalence of latent infection may be explained partly by our model’s assumption that all individuals are equally susceptible to infection (when in reality the existence of “resisters”^24^ may set a ceiling on the prevalence of infection). Finally, the complexity of our agent-based approach has disadvantages that include stochasticity and relatively inefficient calibration – which result in larger uncertainty ranges. However, this complexity enables representation of outcomes in both individual TPT recipients and the general population, interpretation of forthcoming trial results, and exploration of variations in intervention implementation.

In summary, this model-based analysis suggests that, if forthcoming trial results indicate 70% efficacy of delamanid or fluoroquinolone TPT in preventing MDR-TB among contacts, then implementing a package of MDR-TB household contact investigation and TPT could avert one case of MDR-TB for every 12 TPT recipients. Less than 20% of this impact would be directly observed in a clinical trial comparing MDR-TPT to isoniazid or placebo with two-year follow-up. Much of the impact would be attributable to prevented transmission, both as a direct result of TPT and through scaled-up household contact investigation leading to earlier MDR-TB case detection. Guidelines and programmatic decisions regarding implementation and scale-up of MDR-TPT should take these expanded benefits into consideration.

## Data Availability

The simulation model is written in C/C++ and is released under the GPL 2.0 Public License. The source code is available at https://github.com/TB-Modeling/mdrtb. The git repository contains the version controls for previous revisions to this model. Version 2 (saved on Branch TPT-GPL2) was used to produce the results in this manuscript.

## Research in context

### Evidence before this study

We searched Pubmed through 3 February 2023 for tuberculosis AND (“preventive treatment” or “preventive therapy” or “prophylaxis”) AND (“drug resistant” or “mdr” or “model”). Contacts of people with multidrug-resistant tuberculosis (MDR-TB) are at high risk to develop MDR-TB as well. Observational studies suggest that MDR-TB preventive treatment for contacts is safe and effective, although the first clinical trial results from this intervention are not yet published. The small number of modeling analyses of MDR-TB preventive treatment have focused on estimating cost-effectiveness based on impact among the recipients themselves. Although this intervention has been estimated to be cost-effective even without considering transmission effects, accounting for population-wide effects could strengthen the case for its adoption.

### Added value of this study

Our household-structured, agent-based transmission model allowed us to simulate an MDR-TB preventive treatment intervention and to estimate the effects both among the recipients and at a population-level. Specifically, we project the impact among trial’s participants during the follow up period and capture the additional impact which would not be measurable in a clinical trial, reflecting the preventive effects among recipients over the longer term, the downstream effects of preventing transmissions from recipients to the broader population, and the benefits of screening and treating MDR-TB contacts for TB disease during implementation of household contact investigation. We found that accounting for these additional effects of MDR-TB preventive treatment implementation could result in a five-fold increase in the estimated MDR-TB cases prevented, relative to the effects that a typical clinical trial of MDR-TB preventive treatment would directly measure.

### Implications of all the available evidence

If MDR-TB preventive treatment for household contacts is proven to safe and effective in forthcoming trials, the impact and cost-effectiveness of adopting this intervention could be much greater than those trial results alone can measure.

## References

1. Fox GJ, Nhung NV, Sy DN, et al. Household-contact investigation for detection of tuberculosis in Vietnam. New England Journal of Medicine 2018; 378(3): 221–9.

2. Gupta A, Swindells S, Kim S, et al. Feasibility of identifying household contacts of rifampin-and multidrug-resistant tuberculosis cases at high risk of progression to tuberculosis disease. Clinical Infectious Diseases 2020; 70(3): 425–35.

3. Boyd R, Ford N, Padgen P, Cox H. Time to treatment for rifampicin-resistant tuberculosis: systematic review and meta-analysis. The International Journal of Tuberculosis and Lung Disease 2017; 21(11): 1173–80.

4. Thwaites G, Nguyen NV. Linezolid for Drug-Resistant Tuberculosis. New England Journal of Medicine 2022; 387(9): 842–3.

5. World Health Organization (WHO). WHO operational handbook on tuberculosis (Module 1 – Prevention): Tuberculosis preventive treatment Geneva; 2020.

6. Fox GJ, Nguyen CB, Nguyen TA, et al. Levofloxacin versus placebo for the treatment of latent tuberculosis among contacts of patients with multidrug-resistant tuberculosis (the VQUIN MDR trial): a protocol for a randomised controlled trial. BMJ open 2020; 10(1): e033945.

7. Seddon JA, Garcia-Prats AJ, Purchase SE, et al. Levofloxacin versus placebo for the prevention of tuberculosis disease in child contacts of multidrug-resistant tuberculosis: study protocol for a phase III cluster randomised controlled trial (TB-CHAMP). Trials 2018; 19(1): 1–11.

8. ClinicalTrials.gov. Identifier: NCT03568383, Protecting Households On Exposure to Newly Diagnosed Index Multidrug-Resistant Tuberculosis Patients (PHOENIx MDR-TB).2018 https://clinicaltrials.gov/ct2/show/NCT03568383?term=phoenix&cond=tuberculosis&draw=2&rank=12023

9. Velen K, Shingde RV, Ho J, Fox GJ. The effectiveness of contact investigation among contacts of tuberculosis patients: a systematic review and meta-analysis. European Respiratory Journal 2021; 58(6).

10. Blok L, Sahu S, Creswell J, Alba S, Stevens R, Bakker MI. Comparative meta-analysis of tuberculosis contact investigation interventions in eleven high burden countries. PloS one 2015; 10(3): e0119822.

11. Lin PL, Ford CB, Coleman MT, et al. Sterilization of granulomas is common in active and latent tuberculosis despite within-host variability in bacterial killing. Nature medicine 2014; 20(1): 75–9.

12. World Health Organization (WHO). Global Tuberculosis Report. Geneva; 2021.

13. Houben RM, Dodd PJ. The global burden of latent tuberculosis infection: a re-estimation using mathematical modelling. PLoS medicine 2016; 13(10): e1002152.

14. World Health Organization (WHO). National TB Prevalence Survey in India 2019-2021. Geneva; 2022.

15. Martinez L, Shen Y, Mupere E, Kizza A, Hill PC, Whalen CC. Transmission of Mycobacterium tuberculosis in households and the community: a systematic review and meta-analysis. American journal of epidemiology 2017; 185(12): 1327–39.

16. Martinez L, Cords O, Horsburgh CR, et al. The risk of tuberculosis in children after close exposure: a systematic review and individual-participant meta-analysis. The Lancet 2020; 395(10228): 973–84.

17. Fox GJ, Barry SE, Britton WJ, Marks GB. Contact investigation for tuberculosis: a systematic review and meta-analysis. European Respiratory Journal 2013; 41(1): 140–56.

18. Velleca M, Malekinejad M, Miller C, et al. The yield of tuberculosis contact investigation in low-and middle-income settings: a systematic review and meta-analysis. BMC infectious diseases 2021; 21(1): 1–12.

19. Seid G, Alemu A, Dagne B, Sinshaw W, Gumi B. Tuberculosis in household contacts of tuberculosis patients in sub-Saharan African countries: A systematic review and meta-analysis. Journal of clinical tuberculosis and other mycobacterial diseases 2022: 100337.

20. Chiang SS, Brooks MB, Jenkins HE, et al. Concordance of Drug-resistance Profiles Between Persons With Drug-resistant Tuberculosis and Their Household Contacts: A Systematic Review and Meta-analysis. Clinical Infectious Diseases 2021; 73(2): 250–63.

21. Dodd PJ, Mafirakureva N, Seddon JA, McQuaid CF. The global impact of household contact management for children on multidrug-resistant and rifampicin-resistant tuberculosis cases, deaths, and health-system costs in 2019: a modelling study. The Lancet Global Health 2022; 10(7): e1034–44.

22. Kendall EA, Azman AS, Maartens G, et al. Projected population-wide impact of antiretroviral therapy-linked isoniazid preventive therapy in a high-burden setting. AIDS (London, England) 2019; 33(3): 525.

23. Mandal S, Bhatia V, Sharma M, Mandal PP, Arinaminpathy N. The potential impact of preventive therapy against tuberculosis in the WHO South-East Asian Region: a modelling approach. BMC medicine 2020; 18(1): 1–10.

24. Gutierrez J, Kroon EE, Möller M, Stein CM. Phenotype definition for “Resisters” to Mycobacterium tuberculosis infection in the literature—A review and recommendations. Frontiers in Immunology 2021; 12: 619988.

25. Vynnycky E, Fine P. The natural history of tuberculosis: the implications of age-dependent risks of disease and the role of reinfection. Epidemiology & Infection 1997; 119(2): 183–201.

26. Horsburgh Jr CR. Priorities for the treatment of latent tuberculosis infection in the United States. New England Journal of Medicine 2004; 350(20): 2060–7.

27. Dale KD, Trauer JM, Dodd PJ, Houben RM, Denholm JT. Estimating long-term tuberculosis reactivation rates in Australian migrants. Clinical Infectious Diseases 2020; 70(10): 2111–8.

28. Andrews JR, Noubary F, Walensky RP, Cerda R, Losina E, Horsburgh CR. Risk of progression to active tuberculosis following reinfection with Mycobacterium tuberculosis. Clinical infectious diseases 2012; 54(6): 784–91.

29. Salvatore PP, Proaño A, Kendall EA, Gilman RH, Dowdy DW. Linking individual natural history to population outcomes in tuberculosis. The Journal of infectious diseases 2018; 217(1): 112–21.

30. Grandjean L, Gilman RH, Martin L, et al. Transmission of multidrug-resistant and drug-susceptible tuberculosis within households: a prospective cohort study. PLoS medicine 2015; 12(6): e1001843.

31. Borrell S, Gagneux S. Infectiousness, reproductive fitness and evolution of drug-resistant Mycobacterium tuberculosis [State of the art]. The International Journal of Tuberculosis and Lung Disease 2009; 13(12): 1456–66.

32. Starke JR. Transmission of Mycobacterium tuberculosis to and from children and adolescents. Seminars in Pediatric Infectious Diseases; 2001: Elsevier; 2001. p. 115–23.

33. Tiemersma EW, van der Werf MJ, Borgdorff MW, Williams BG, Nagelkerke NJ. Natural history of tuberculosis: duration and fatality of untreated pulmonary tuberculosis in HIV negative patients: a systematic review. PloS one 2011; 6(4): e17601.

34. Ragonnet R, Flegg JA, Brilleman SL, et al. Revisiting the natural history of pulmonary tuberculosis: a bayesian estimation of natural recovery and mortality rates. BioRxiv 2019: 729426.

35. Subbaraman R, Nathavitharana RR, Satyanarayana S, et al. The tuberculosis cascade of care in India’s public sector: a systematic review and meta-analysis. PLoS medicine 2016; 13(10): e1002149.

36. MacPherson P, Houben RM, Glynn JR, Corbett EL, Kranzer K. Pre-treatment loss to follow-up in tuberculosis patients in low-and lower-middle-income countries and high-burden countries: a systematic review and meta-analysis. Bulletin of the World Health Organization 2013; 92: 126–38.

37. World Health Organization (WHO). Global Tuberculosis Report. Geneva; 2019.

38. Ahuja SD, Ashkin D, Avendano M, et al. Multidrug resistant pulmonary tuberculosis treatment regimens and patient outcomes: an individual patient data meta-analysis of 9,153 patients. PLoS medicine 2012; 9(8): e1001300.

39. O’Donnell MR, Padayatchi N, Kvasnovsky C, Werner L, Master I, Horsburgh Jr CR. Treatment outcomes for extensively drug-resistant tuberculosis and HIV co-infection. Emerging infectious diseases 2013; 19(3): 416.

40. Marx FM, Dunbar R, Enarson DA, et al. The temporal dynamics of relapse and reinfection tuberculosis after successful treatment: a retrospective cohort study. Clinical infectious diseases 2014; 58(12): 1676–83.

41. Menzies D, Benedetti A, Paydar A, et al. Effect of duration and intermittency of rifampin on tuberculosis treatment outcomes: a systematic review and meta-analysis. PLoS medicine 2009; 6(9): e1000146.

42. Menzies D, Benedetti A, Paydar A, et al. Standardized treatment of active tuberculosis in patients with previous treatment and/or with mono-resistance to isoniazid: a systematic review and meta-analysis. PLoS medicine 2009; 6(9): e1000150.

