## Supplement for "Trials underestimate the impact of preventive treatment for household contacts exposed to multidrug-resistant tuberculosis: a simulation study"

Kasaie et al.

### 1 SIMULATION MODEL ARCHITECTURE

#### 1.1 Population design

The baseline simulation models a self-contained population, representing a hypothetical city in India. Simulated individuals enter the population at birth and leave the model upon death. Each simulated agent ages out of the model at 85 years, and those with active TB are subject to an additional risk of TB mortality.

Population growth is modeled on an annual basis by introducing a cohort of newborns into the model at the start of each year. Using data on the reported and projected population size of India from the United Nations Department of Economic and Social Affairs [1] (**Figure S1-A**), we estimated the annual population growth rate for years between 2000 to 2040. For years before 2000, we assumed a fixed annual growth rate at the value for year 2000 (**Figure S1-B**).

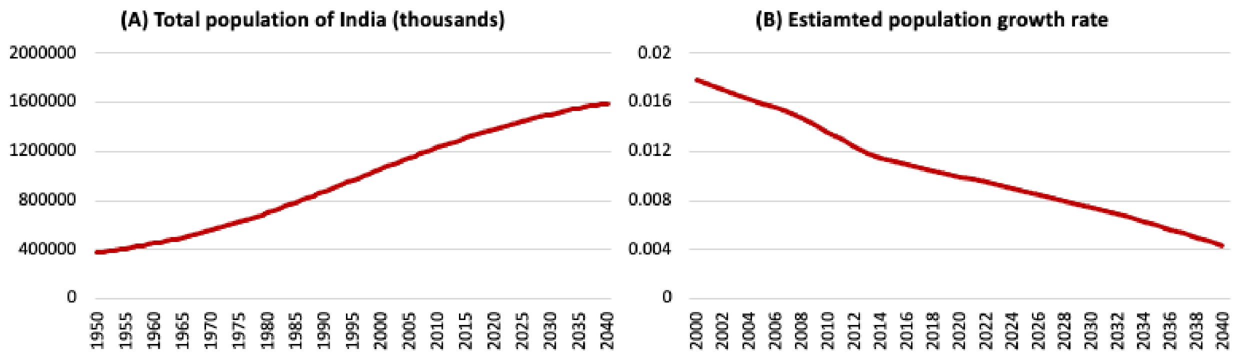

**Figure S1: Population size and growth rate projections.** Panel A presents the reported total population (both sexes combined) of India from 1950-2020 as of July 1 of the year indicated and the projected total population from 2021-2040 assuming a medium fertility variant. Panel B presents the estimated annual population growth rate from 2000-2040.

#### 1.2 Household design

We adopted a simplified household structure that would provide a realistic representation of TB transmission dynamics among household members by age and sex. To this end, we estimated the empirical distribution of household sizes, as well as age/sex composition of household members by household size, from a representative survey of India's households.[2] For computational efficacy, we only focused on modeling the households of individuals with current or recent active TB, and followed these households for a limited duration of time that would allow us to model most opportunities for household TB transmission or household-targeted TB prevention (see below).

##### 1.2.1 Household creation

We used data from the India Human Development Survey-II (IHDS-II) [2] to estimate the empirical distribution of household sizes, and sex/age composition, for simulated agents in our model. IHDS-II a is a national survey of 42,152 households in 1,503 villages and 971 urban neighborhoods across India, covering topics related to health, education, employment, economic status, marriage, fertility, etc. We extracted information on surveyed households' size (ranging from 1 to 33 people), as well as the number of male and female children (<14y), teens (14-20y) and adults (>20) within each household.

In the model, the "creation" of a new household is triggered by the onset of active TB (ATB), unless the incident case already belongs to an existing household. In reality, households exist before the onset of active TB, but this mechanism enables the model to begin following specific households at the time of onset of TB disease without the need to continuously represent all households (and their corresponding dynamics) in the underlying population. To "create" a new household for a person with incident TB (say, for illustrative purposes, a 33y woman with drug-susceptible [DS] ATB), we first filter the household survey dataset to consider only households with at least one member within the index case's sex/age category (e.g., households with at least one female adult). In the next step, a random household is sampled from the list of available households (which might, for instance, result in sampling of a household of size five with two female adults, one male adult, and two male children). In preparation for selecting the other members of the new household, we then sort the available population members (those not currently assigned to any household) into demographic bins. After assigning the index case to the newly created household, the remaining household members are selected randomly from their respective demographic bins. If no eligible individual is found within a bin, a smaller household is formed which omits that member.

##### 1.2.2 Household follow up and dissolution

Each household is followed for a limited period before dissolution. This period is set to 10 years in our baseline model, which is long enough to capture the household linkage of most secondary TB cases that result from transmission from the index case to other household members and is short enough to allow for realistic dissolution of households due to population dynamics (e.g., marriages resulting in divorces, kids growing up and leaving home, etc). The follow-up period is extended if any household members develop active TB (e.g., if a new household member develops active TB after five years of follow up, the household will be followed for another 10 years – for a total follow-up time of 15 years). The follow up period is also extended until no household members have active disease. Upon reaching the end of the follow up period, the household is dissolved, and household members are returned to the general community. These individuals will then be eligible for assignment to new households.

##### 1.2.3 Model initialization and annual dynamics

Given the slow dynamics of TB epidemics, simulations are initialized with a population of 100,000 individuals at a historical time representative of the year 1500. The DS-TB epidemic is seeded by 50 active TB cases, chosen randomly among the initial population. The first cohort of households is created for these active TB cases, and household dynamics (as described above) are modeled on a monthly basis.

Population demographics are updated on an annual basis by modeling deaths (e.g., natural death, aging out at the age of 85, and/or TB mortality), and new births (e.g., assuming a population growth rate at 1.7% pre-2000). A specific proportion of newborns (set to the proportion of the population that are in households at the beginning of each simulated year) are assigned to randomly selected households, and the remainder of newborns enter the general community.

To maintain a computationally tractable size for the simulated population, we implemented a "population stabilization" process (described below) to keep the population size roughly constant near 100,000 individuals for the first 400 years of simulation from 1500 – 1900. This initial period allowed us to reach DS-TB equilibrium. Starting in year 1900, we discontinued the stabilization process and allowed the simulated population size to increase through year 2040 according to the estimated population growth rates in **Figure S1-B**, reaching approximately 800 thousands by 2022. All individual trajectories were followed until death or year 2040.

##### 1.2.4 The "Population Stabilization" Process

The stabilization process is implemented every 20 years, by removing a random sample of individuals and households from the population. As described below, the model is structured such that only a subset of the individuals in the populations are assigned to a household at any given time. Thus, the stabilization process proceeds as follows:

**1) Estimating the proportion of individuals that should be removed ($P^{removal}$)**

$P^{removal} = 1 - N0/N(t)$ where $N(t)$ is the current population size at time $t$ and $N0$ is the target population size at 100,000 individuals.

**2) Determining the number of household-free individuals to remove ($N^{removal\text{-}HH\text{-}free}$)**

$$N^{removal\text{-}HH\text{-}free} = P^{removal} * N^{HH\text{-}free}(t)$$

where $N^{HH\text{-}free}(t)$ is the number of individuals with no household assignment ("household-free individuals") at time $t$,

**3) Determining the number of households to remove ($N^{removal\text{-}HHs}$)**

$$N^{removal\text{-}HHs} = P^{removal} * N^{HHs}(t)$$

where $N^{HHs}(t)$ is the number of households at time $t$,

**4) Removal:** From the list of household-free individuals, randomly select and remove $N^{removal\text{-}HH\text{-}free}$ individuals from the population. From the list of households, randomly select and remove $N^{removal\text{-}HHs}$ households (including all members of those households) from the population.

#### 1.3 Contact network

To capture the heterogeneous pattern of TB transmission within households and the community, we model two types of contact events: 1) "close" contact (with household members), and 2) "casual" contact (with other members of the community). Age-dependent frequencies of casual contacts in each month are estimated from literature.[3] In addition, each contact type is associated with an crude ATB transmission risk parameter describing the force of TB transmission, allowing us to model a higher risk of transmission via close compared to casual contact.

**Data source:** The frequency of casual contact is estimated based on Prem et al. (2017), which estimates the age-specific frequency of daily contacts with household and non-household (e.g., work, school, other) community members.[3] Using data from a previous population-survey of eight European countries (POLYMOD study), we applied a Bayesian hierarchical model to project age-and-location-specific contact patterns for 144 other countries including India.[4] We used this data to estimate the monthly frequency of non-household contacts, and the proportion of contacts that occur between various age-groups (**Table S1**).

**Table S1: Frequency and proportion of casual contacts between different age groups**

| Age (y) | Number of monthly contacts (N) | Proportion of contacts within each age-group | | | | | | | | | | | | | | | |
|---|---|---|---|---|---|---|---|---|---|---|---|---|---|---|---|---|---|
| | | [0,5) | [5,10) | [10,15) | [15,20) | [20,25) | [25,30) | [30,35) | [35,40) | [40,45) | [45,50) | [50,55) | [55,60) | [60,65) | [65,70) | [70,75) | [75,85) |
| **[0,5)** | 216 | 0.28 | 0.111 | 0.056 | 0.042 | 0.066 | 0.09 | 0.097 | 0.077 | 0.048 | 0.033 | 0.034 | 0.026 | 0.016 | 0.013 | 0.008 | 0.003 |
| **[5,10)** | 491 | 0.054 | 0.617 | 0.092 | 0.023 | 0.018 | 0.034 | 0.04 | 0.039 | 0.032 | 0.015 | 0.011 | 0.009 | 0.007 | 0.005 | 0.002 | 0.001 |
| **[10,15)** | 542 | 0.015 | 0.146 | 0.584 | 0.057 | 0.031 | 0.025 | 0.028 | 0.031 | 0.034 | 0.02 | 0.012 | 0.006 | 0.004 | 0.004 | 0.002 | 0.002 |
| **[15,20)** | 710 | 0.007 | 0.023 | 0.168 | 0.551 | 0.084 | 0.038 | 0.027 | 0.029 | 0.028 | 0.023 | 0.011 | 0.005 | 0.003 | 0.002 | 0.001 | 0.001 |
| **[20,25)** | 467 | 0.013 | 0.019 | 0.028 | 0.23 | 0.311 | 0.12 | 0.076 | 0.061 | 0.045 | 0.042 | 0.027 | 0.015 | 0.006 | 0.002 | 0.002 | 0.002 |
| **[25,30)** | 362 | 0.025 | 0.025 | 0.018 | 0.079 | 0.188 | 0.221 | 0.126 | 0.099 | 0.078 | 0.058 | 0.046 | 0.023 | 0.009 | 0.003 | 0.001 | 0.001 |
| **[30,35)** | 320 | 0.028 | 0.061 | 0.055 | 0.048 | 0.096 | 0.138 | 0.168 | 0.128 | 0.098 | 0.074 | 0.053 | 0.033 | 0.013 | 0.004 | 0.002 | 0.002 |
| **[35,40)** | 297 | 0.026 | 0.054 | 0.039 | 0.046 | 0.068 | 0.118 | 0.134 | 0.166 | 0.139 | 0.089 | 0.062 | 0.031 | 0.015 | 0.007 | 0.004 | 0.001 |
| **[40,45)** | 279 | 0.017 | 0.036 | 0.046 | 0.074 | 0.075 | 0.101 | 0.125 | 0.132 | 0.155 | 0.108 | 0.078 | 0.029 | 0.015 | 0.005 | 0.003 | 0.001 |
| **[45,50)** | 224 | 0.014 | 0.047 | 0.037 | 0.116 | 0.062 | 0.092 | 0.118 | 0.127 | 0.126 | 0.117 | 0.081 | 0.04 | 0.014 | 0.004 | 0.003 | 0.002 |
| **[50,55)** | 249 | 0.009 | 0.063 | 0.07 | 0.093 | 0.065 | 0.097 | 0.096 | 0.097 | 0.123 | 0.121 | 0.091 | 0.05 | 0.017 | 0.005 | 0.003 | 0.002 |
| **[55,60)** | 185 | 0.022 | 0.07 | 0.065 | 0.077 | 0.058 | 0.101 | 0.112 | 0.101 | 0.116 | 0.085 | 0.089 | 0.065 | 0.027 | 0.008 | 0.004 | 0.002 |
| **[60,65)** | 106 | 0.023 | 0.035 | 0.031 | 0.067 | 0.068 | 0.098 | 0.105 | 0.129 | 0.113 | 0.1 | 0.08 | 0.072 | 0.041 | 0.022 | 0.012 | 0.005 |
| **[65,70)** | 46 | 0.03 | 0.061 | 0.033 | 0.027 | 0.075 | 0.101 | 0.125 | 0.1 | 0.088 | 0.067 | 0.062 | 0.072 | 0.059 | 0.059 | 0.031 | 0.013 |
| **[70,75)** | 42 | 0.011 | 0.024 | 0.036 | 0.073 | 0.058 | 0.074 | 0.07 | 0.095 | 0.103 | 0.082 | 0.074 | 0.062 | 0.076 | 0.069 | 0.061 | 0.033 |
| **[75,85)** | 20 | 0.032 | 0.065 | 0.051 | 0.06 | 0.058 | 0.078 | 0.117 | 0.087 | 0.076 | 0.086 | 0.057 | 0.051 | 0.045 | 0.06 | 0.049 | 0.03 |

**Simulation process:** At the beginning of each simulated month, we model two types of contacts for infectious individuals (those with active TB or failing treatment): 1) close contact with all household members, 2) casual contacts with a random selection of community members. The number of casual contacts (N) for an active TB case is determined based on each agent's age group (second column in **Table S1**). We then determine the number of contacts with individuals in each age group by multiplying the number of contacts (N) by the various proportions (rounding up). Subsequent partners are selected randomly from community members in each age group with no replacement. If no individual is found within an age group, the corresponding age-specific casual contacts are omitted.

##### 1.4 TB natural history

At an individual level, the natural history of TB is modeled as shown in **Figure 1** in the main text. We model circulation of both drug-susceptible (DS) and rifampin-resistant (MDR) TB strains in the population.

#### 1.4.1 Progression to active disease

Each person is born in full health and susceptible to TB disease. When successful transmission of TB infection occurs, the infected person enters the Early Latent TB state for a period of five years, during which active TB may develop via *primary progression.*[5] The monthly probability of primary progression decreases over time during these first five years ($Pr(t), t = 1, \dots ,60$ ,), as follows:

$$Pr(t) = cFP_0 \times FP_{age} \times rrFP(t) \,; t = 1, \dots ,60$$

Where $cFP_0$ is a calibrated coefficient describing the five-year cumulative risk of primary progression among persons >15 years old at the time of infection (**Table 1**), $FP_{age}$ is the relative age-specific five-year cumulative risk of primary progression compared to persons >15 years old (**Table 1**), and $rrFP(t)$ is an exponential decay function describing how the risk is distributed over time during first 60 months post infection:

$rrFP(t) = Q/\sum_{t=0}^{60} Q$, where $Q = e^{-0.075t}$ is a function calibrated to annual cumulative probabilities (relative to year 1) of 0.41 in year 2, 0.13 in year 3, 0.086 in year 4, and 0.028 in year 5 [5], and *t* is time since infection in months.

At the end of this five-year period, if the person has not developed active TB or been reinfected with the same strain, they enter the Late Latent TB state, which can last for many years and is associated with a lower and constant probability of progression to active TB via *reactivation*.

#### 1.4.2 Pediatric TB

In order to capture the impacts of household contact interventions on TB morbidity and mortality in young children, the TB natural history described above is age-dependent. Young children (infected at age 0-2 years) have a higher risk of primary progression from early latent to active TB compared to adults, while older children (infected at age 2-10 years) have lower risk of primary progression than adults (**Table 1**). Children who do progress to active TB before 10 years of age are considered less infectious than adults with TB disease, although they face the same mortality risk as other ATB. In late latent infection, children are assumed to have the same monthly risk of reactivation as adults. Infections acquired during childhood that progress to active TB after age >10 years are as infectious as other adult TB cases.

#### 1.4.3 Active TB infectivity and outcomes

For the first nine months of disease, the infectivity of individuals with active TB is modeled as increasing linearly over time, from zero at the start of disease to a peak level nine months later; it remains constant at the peak level thereafter for the duration of untreated disease.[6] The peak infectiousness of MDR-TB (per infectious person-week) is allowed to be lower than that of DS-TB.

Active TB can end in outcomes of death, spontaneous resolution, or successful treatment. Individuals with active TB experience an increased risk of mortality, which is additive to their natural age-related mortality and rises to a plateau over the first nine months of disease in the same linear fashion as infectivity. Individuals with active TB can recover from disease through spontaneous resolution, modeled as a constant probability per time step during time periods with untreated disease. Finally, to recover through treatment, patients must present to care and start a successful treatment regimen. Recovery occurs after the full duration of treatment has passed, but patients are noninfectious during successful treatment. During unsuccessful treatment (i.e., treatment which will end in failure and continued active TB), infectivity continues at a reduced level.

#### 1.4.4 TB diagnosis and treatment

The monthly probability of seeking care and being evaluated for TB increases over the first nine months of disease, in the same linear fashion as infectivity and mortality. Upon presentation to care, all patients with presumptive TB are assumed to receive a bacteriologic test to diagnose DS-TB infection (i.e., if such a test was not performed during a visit, that visit would not be considered in our analysis). A proportion of patients will also receive a drug susceptibility test (DST) to detect MDR-TB; the probability of this DST changes over time (reflecting changes in availability) and may depend on their treatment history.

**DST availability:** Using WHO's reported data on TB notification in India, [7] we estimated the proportion of new and relapse TB cases with MDR-TB disease that were notified as MDR-TB in each year. We observed an approximately linear increase in the number of MDR-TB diagnoses over time, from near 0 in 2008, to approximately 25% of our estimate of the number of actual MDR-TB cases among new and relapse TB cases in 2016. Meanwhile, DST coverage was reported to have reached ~60% among retreatment patients by 2014, and to have begun to scale up among new patients in 2014. The rate of increase in new DST coverage after 2014 was fit to the observed trends in overall MDR-TB notifications, assuming separate linear rates of increase in coverage proportion among new and retreatment patients. As such, we simulated the probability of DST among new and retreated patients over the simulated years as follows:

1) Linear increase in DST coverage from 0% to 60% (10%/year increase) among retreatment patients from 2008 to 2014, followed by linear increase to reach 100% coverage by 2024 (**Figure S2-orange line**).

2) Linear increase in DST coverage among new patients from 0 in 2014 to 30% in 2017 (a 10%/year linear scaleup), followed by linear scale up to reach 100% coverage by 2024 (**Figure S2-blue line**).

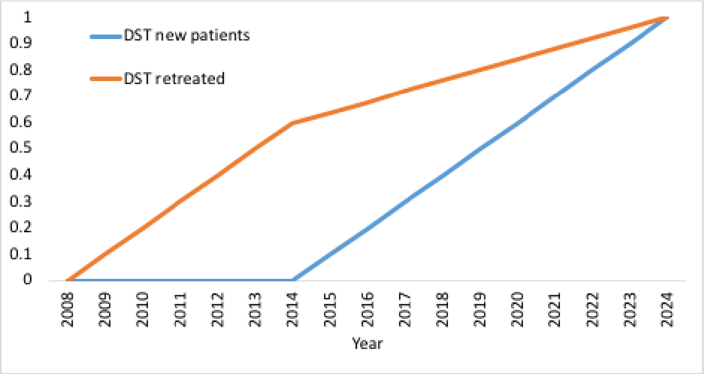

**Figure S2: Estimate probability of DST for new and retreated patients over time.**

Upon TB diagnosis, patients experience a one-time probability of pre-treatment loss to follow up (LTFU) (**Table 1**). Those who are lost to follow up are assumed to continue with active disease but can return to seek care at a future time. The remainder of patients are assumed to receive treatment according to their diagnosis. All patients who initiate TB treatment may be cured, die of TB, or fail treatment.

We assume that all individuals initiating treatment for active TB are initiated on a 6-month, four-drug treatment regimen for DS-TB unless they receive an DST result indicating resistance to rifampin – in which case we assume that they are initiated appropriately on 20-month treatment for MDR-TB. We further assume that DS-treatment has no effect against latent DR-infection and can only clear 10% of active DR-infections. On the other hand, DR-treatment can clear existing DS-TB infections even if it fails in clearing the active MDR-TB disease (e.g., a patient with active MDR-TB and latent DS-TB who fails DR-treatment will return to the active MDR-TB state but recover from latent DS-TB). While we acknowledge that shorter regimens for MDR-TB are likely to become standard in future years, we do not explicitly incorporate shorter MDR-TB regimens in this analysis, for purposes of simplicity and transparency (and so as not to make explicit assumptions about the availability of such regimens in resource-constrained settings).

##### 1.4.5 Treatment failure and evolution of drug-resistance

All patients receiving treatment for active TB experience a probability of treatment failure, upon which they remain partially infectious and continue to experience a risk of TB mortality throughout the non-curative treatment period.

Upon the end of non-curative treatment, individuals return to the active disease state and can return to care for future diagnostic attempts.

Additionally, we model a similar mechanism for DS-TB treatment failure that leads to acquiring drug resistance. In this case, the patient recovers from DS-TB (moving to DS-REC state, **Figure 3** in the main text), but they subsequently develop active MDR-TB disease after the treatment is completed. The onset of MDR-TB disease is set at the end of failed DS-TB treatment, followed by 9 months of increasing severity as for new incident disease. These patients remain infectious with MDR-TB until death, spontaneous resolution, or initiation of successful treatment occurs.

Upon presentation to care, MDR-TB patients may receive DST, with a probability that depends on the calendar year and their treatment history as described above. In absence of DST, MDR-TB patients will be misdiagnosed with DS-TB, and they will initiate a course of DS-TB treatment that fails in the majority of cases. During the non-curative treatment period, the patients remain partially infectious with MDR-TB, and they continue to experience a risk of TB mortality. Upon completion of failed treatment, these patients resume care-seeking and can be diagnosed and offered treatment again.

##### 1.4.6 Mixed infections

All individuals in latent or recovered TB states are subject to reinfection with either strain. To preserve a record of MDR-TB risk when a person with latent MDR-TB experiences DS-TB infected and/or treatment, we allow simultaneous latent infections with both DS-TB and MDR-TB. For each resistance profile (DS and MDR), we only allow a single infection, modeled after the trajectory of the most recent infection with that resistance profile. For simplicity, once active TB occurs with either strain, we do not allow progression to active disease from the other strain until the original episode of active disease ends, such that dual active disease does not occur (e.g., an individual may develop active MDR-TB and be infected with latent DS-TB at the same time; but cannot develop active MDR-TB and active DS-TB simultaneously).

##### 1.4.7 Immunity toward (re)infection

A history of prior or current TB infection may offer a degree of immunity against additional infection with either strain (**Table 1**). This is modeled as a coefficient of immunity and is modeled independent of the TB strain (i.e., any existing infection with either strain offers immunity toward future infection with both strains). We assume partial immunity toward reinfection in the latent and recovered states. We also use the same coefficient to prevent additional infections during active disease and while on TB treatment.

##### 1.4.8 Representing dynamic changes in TB epidemiology

We used a combination of two mechanisms to replicate global reductions in TB infections in 21st century: reductions in TB transmission probabilities and reductions in TB reactivation rates. In the absence of direct estimates, we modeled each of these effects as a rate of change after year 2000, each with a uniform prior distribution with a range from 0 to an upper bound empirically chosen to exceed the maximal calibrated value (**Table 1**).

##### 1.4.9 TB transmissions

TB transmission can occur via household and/or non-household contacts of active TB patients. New infections are updated at the end of each simulated month. The probability of transmission $Ptran(p,q,c,t)$ from an infectious individual $(p)$ to a contact $(q)$ with whom they have contact of type $(c)$ at time $t$ is calculated as follows:

$$Ptran(p,q,c,t) = Inf(p,t) \times Imm(q,t) \times Int_{(close,casual)} \times Red.trans_{post-2000}(t)$$

$$Inf(p,t) = \begin{cases} \frac{\lfloor t - t_0 \rfloor}{9} \times MaxInf & if\ (t - t_0) \leq 9 \\ MaxInf & o.w. \end{cases}$$

$Inf(p,t)$ represents the infectivity of person $p$ at month $t$, and is modeled as a step function with linear monthly increase over the first 9 months after disease onset (month $t_0$) before reaching the maximum level of infectiousness ($MaxInf$) and staying at that level afterward. In addition, $Imm(q,t)$ represents the immunity of person $q$ toward infection/reinfection, based on that individual's prior infection status (**Table 1**). Finally, $Int_{close}$ & $Int_{casual}$ represent the crude TB transmission risk via close contacts with household members or casual contacts with community members, and $Red.trans_{post-2000}(t)$ represents the modeled reduction in probability of transmission after the year 2000.

#### 1.5 Simulation calibration

The model calibration process proceeds in 2 steps as shown in **Figure S3**. In Stage 1, we focus on calibrating DS-TB dynamics to the year 1970 before introducing MDR-TB infections, and in Stage 2, we focus on calibrating DS-TB and MDR-TB dynamics to 2022 (simulation baseline year) before modeling new interventions (2023 forward). At the end of Stage 2, a Sampling Importance Resampling (SIR) approach is used to construct a posterior distribution of simulations consistent with available data.

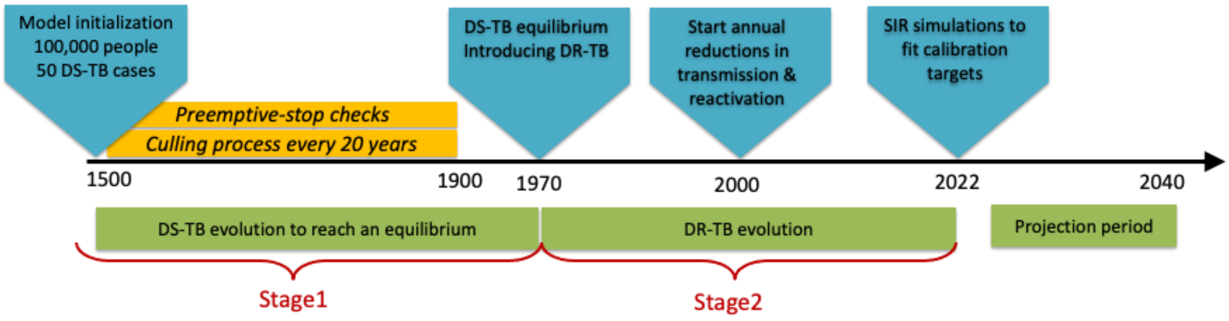

**Figure S3: Simulation calibration stages.**

##### 1.5.1 Simulation Stage 1

Prior distributions were defined for all simulation parameters corresponding to TB transmission and DS- and DR-TB natural history (**Table S2**). Priors were informed by published data when available, and otherwise, we used minimally informative prior distributions and checked manually that the posterior had low density at the bounds of the sampled interval (widening the prior when it did not).

In Stage 1 of parameter sampling, we drew 10,000 samples of 13 DS-related parameters from their corresponding prior distributions using Latin Hypercube Sampling. For each sampled parameter set, we ran a randomly seeded simulation from year 1500 to year 1970 (unless early stopping criteria were met, described below).

**Preemptive-stop checks:** To save computational time, we established early stopping criteria ("preemptive-stop checks") to filter out simulation runs that followed an unrealistic trajectory and were highly unlikely to be selected in importance resampling. This is particularly important for simulations that generate an unrealistically high TB incidence, because simulations run most slowly when a large proportion of the population has active TB (as might result, for example, when high values of all transmission and progression parameters are simultaneously sampled). Therefore, we periodically checked the simulated rate of DS-TB incidence against an upper threshold of 500 cases per 100,000 people (approximately 4 standard deviations above the target incidence for year 2000) and aborted simulations that fell above this threshold. This check occurred every 10 years between year 1500 to 1600, and every 100 years between year 1600 to 1900. In 1900, we also checked the DS-TB incidence rate against a lower threshold of 100 cases per 100,000 (approximately 4 standard deviations below the target incidence for year 2000), aborting simulations that fell below this threshold. Following this procedure, 9062 simulations (out of 10,000 Stage1 simulations) failed the preemptive-stop check and were aborted. 938 simulations passing these preemptive-stop checks continued to year 1970.

**Table S2: List of model parameters and prior distributions in each calibration stage**

| **Stage 1 parameters** | **Prior distribution** | **Reference, Notes** |
|---|---|---|
| Infectiousness of children £10y, relative to adults | BETA [2.6E-05, 1.0E-03, 5.8E-03] | [8] |
| Probability of pretreatment loss-to-follow-up, DS-TB | BETA [0.10, 0.13, 0.17] | [9,10] |
| Annual probability of reactivation >5 years post infection | BETA [5.4E-04, 7.5E-04, 1.0E-03] | [11] [12,13] |
| Monthly probability of spontaneous resolution from ATB | BETA [2.31E-03, 0.01, 0.02] | [14,15] |
| Relative reduction in reinfection risk if latently infected | BETA [0.14, 0.30, 0.50] | [16] [5] |
| Probability of relapse in the two years after resolution, DS-TB | BETA [0.04, 0.06, 0.09] | [17] [18] |
| Cumulative probability of primary TB disease in the first 5 years post infection among persons >15 years old at the time of infection | R= LOGNORM[0.06, 0.09, 0.13] | [5] |
| Maximum monthly probability of ATB mortality | UNIFORM [0.0 – 0.03] | Assumption |
| Infectiousness while failing treatment, relative to untreated TB | UNIFORM [0 – 1] | |
| Crude TB transmission risk among household contacts (per month) | UNIFORM [0.0 – 0.3] | |
| Crude TB transmission risk among non-household contacts (per month) | UNIFORM [0.0 – 0.005] | |
| Maximum monthly probability of seeking care for ATB | UNIFORM [0.1 – 0.15] | |
| Probability of treatment failure, DS-TB | BETA [0.02, 0.03, 0.04] | [19] |
| **Stage 2 parameters** | | |
| Probability of pretreatment loss-to-follow-up, MDR-TB | BETA [0.18, 0.23, 0.29] | [9,10] |
| Probability of treatment failure, MDR-TB (appropriately treated) | BETA [0.17, 0.24, 0.32] | [18] |
| Probability of treatment failure, MDR-TB receiving DS-TB regimen | BETA [0.78, 0.90, 0.97] | [20] |
| Probability of relapse in the two years after resolution, MDR-TB | BETA [0.04, 0.06, 0.09] | [17] [18] |
| Infectiousness of MDR-TB, relative to DS-TB | BETA [0.41, 0.75, 0.95] | [21-23] |
| Probability of acquiring resistance following DS-TB treatment (after 1970) | BETA [9.05E-04, 5.00E-03, 0.02] | [24,25] |
| Annual reduction in TB transmission post-2000 | UNIFORM [0.0 – 0.012] | Assumption |
| Annual reduction in TB reactivation post-2000 | UNIFORM [0.0 – 0.012] | |

##### 1.5.2 Simulation Stage 2

Starting in year 1970, MDR-TB infections were introduced to the model by allowing a nonzero proportion of those who complete DS-TB treatment to acquire resistance. In this stage, we sampled eight additional parameters that had not been relevant in Stage 1 of the simulation, related to MDR-TB dynamics or the decline in TB incidence after year 2000. For each simulation that reaches 1970 in Stage 1, these eight parameters were sampled via Latin Hypercube Sampling with 100 rows (LHC-100, unique for each pre-1970 simulation run), resulting in 100 nested simulations beyond year 1970.

Following simulation stage 1, each of the 938 resulting simulations (each of which had been saved in year 1970 at the end of stage 1) was replicated into 100 parallel simulations and run from 1970 to 2021 for 100 unique sets of sampled values for the Stage 2 parameters. For this stage, we performed a single preemptive-stop check in year 2000: the proportion of retreatment TB cases that have MDR-TB (whether it is diagnosed as such) was calculated as an average over the preceding 5 years (1995-1999), and runs were aborted if this proportion exceeded a threshold of 40%. Following this procedure, 46,669 simulations (out of 93,800 Stage 1 simulations) failed the preemptive-stop check in Stage 2 and were aborted. For models that proceed past this check (47,131 remaining models), the model state was saved in year 2022.

##### 1.5.3 Model fitting

A Bayesian approach was used to construct a posterior set of simulations that reflect available knowledge about input parameter values and about epidemiological data used as calibration targets (**Table S3**). As part of this process, a psudo-likelihood was calculated for each model at the end of simulation Stage 2. Uncertainty in each calibration target $j$ was represented as a log-normal or logit-normal distribution (for outcomes that are proportions or have no theoretical upper bound, respectively), with mean $m_j$ based on available point estimates and variance based on the width of the 95% confidence interval for the target estimate ($lb_j$, $hb_j$) after log- or logit transformation. The corresponding model outputs $sim_j^i$ for simulation $i$=1,…,$N$ were compared to these targets to calculate a psudo-likelihood for target $j$. Because relatively orthogonal outcomes were chosen as calibration targets (e.g. incidence and average duration, rather than incidence and prevalence), a joint psudo-likelihood $l$ was calculated as a product of individual psudo-likelihoods $l_i$. psudo-log-likelihoods $ll$ = log($l$) were used for computational convenience. The joint psudo-log-likelihood was thus calculated as follows:

$$ll(i) = \sum_{j=1}^{8} ll_j^i \quad i = 1, \dots, N$$

$$ll_j^i = \log\left( dnorm\left( \log(sim_j^i) - \frac{\log(m_j)}{(\log(hb_j) - \log(lb_j))/4}\right)\right) \; if \;\; j \in \mathbb{R}$$

Or

$$ll_j^i = \log\left( dnorm\left( \text{logit}(sim_j^i) - \frac{\text{logit}(m_j)}{(\text{logit}(hb_j) - \text{logit}(lb_j))/4}\right)\right) \; if \; j \in [0, \dots, 1]$$

**Table S3: Calibration targets and values**

| Calibration target | Med | LB | UB | Reference |
|---|---|---|---|---|
| TB incidence per 100,000 persons in 2019 | 193 | 132 | 266 | [26] |
| TB incidence in 2019, relative to 2000 | 0.67 | 0.46 | 0.92 | 2% annual reduction; [26] |
| TB mortality per 100,000 persons in 2019 | 33 | 16 | 50 | [26] allowing ±50% uncertainty given limitations of mortality data |
| Proportion of newly diagnosed TB patients that have MDR-TB in 2019 | 0.03 | 0.02 | 0.04 | [26] |
| Proportion of retreated TB patients that have MDR-TB in 2019 | 0.14 | 0.12 | 0.16 | [26] |
| Prevalence of latent TB in 2019 | 0.33 | 0.16 | 0.50 | [27] allowing ±50% uncertainty to account for upward revision of India’s TB burden estimates but declining prevalence over time, since LTBI prevalence was estimated |
| Proportion of incident TB that results from household transmission | 0.2 | 0.1 | 0.4 | [28] with upper bound increased to account for potential transmission from household source cases who were never diagnosed |
| Prevalence of TB per 100,000 persons 2019 | 316 | 290 | 342 | Adjusted prevalence of TB for all age-groups from 2019 to 2021 (Table 21 in [29]) |

##### 1.5.4 Likelihood estimates

The likelihood was calculated for each of these 47,131 models that successfully reached the end of Simulation Stage 2. Among these simulations, 555 models accounted for 99.99% of the weighted joint likelihood. **Figure S4** shows the fit to the main calibration targets among these 555 simulations. **Figure S5** compares the prior vs posterior distributions among these runs.

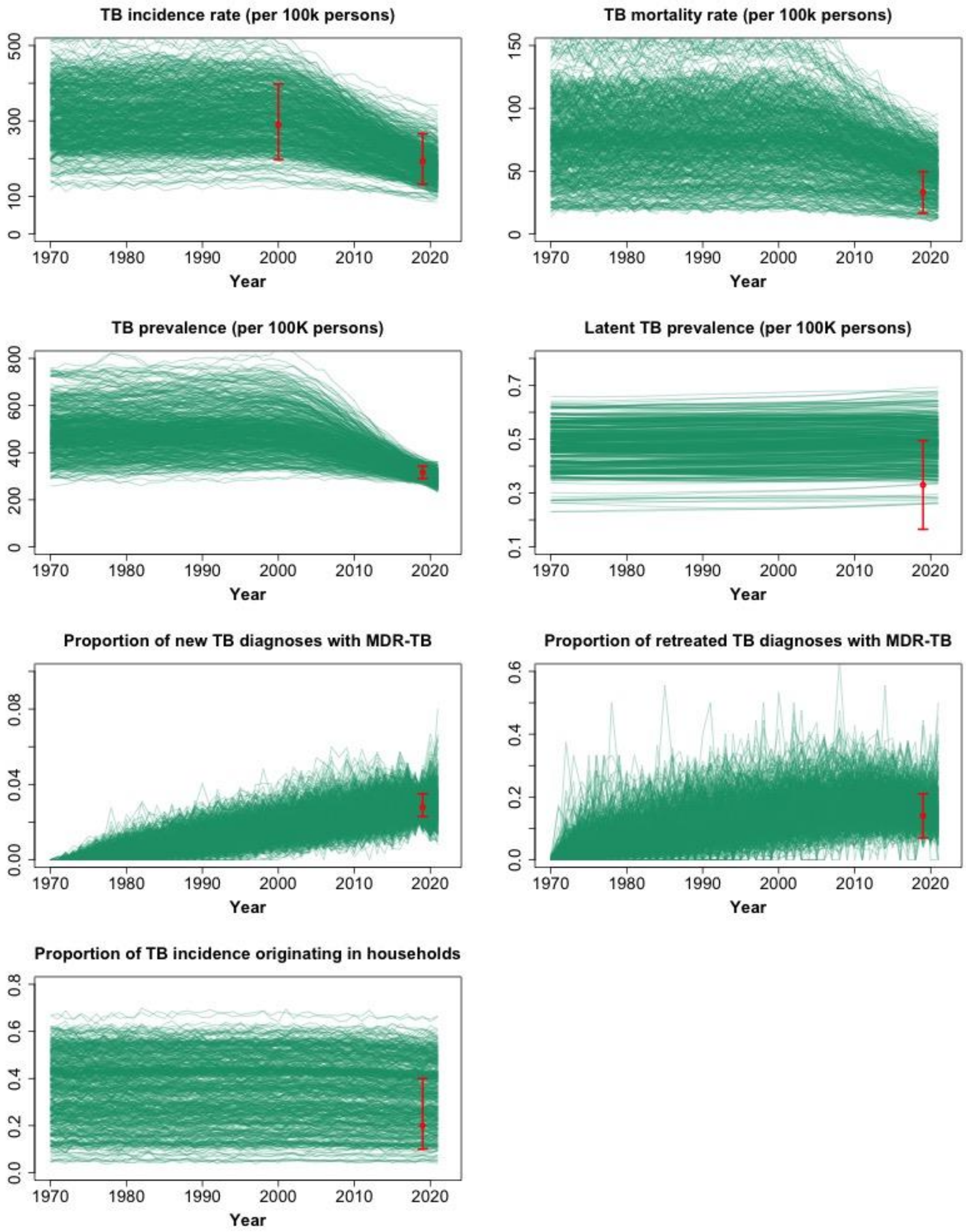

**Figure S4: calibration summary**. Panels represent individual fit to main calibration targets among the top 555 simulations during the calibration period, corresponding to the 99.99% of the weighted joint psudo-likelihood. Red dots represent the median of each calibration target that contributed to the likelihood, and the red error bars represent the outer 0.95 quantile range of each calibration target. Using the likelihood values as weights, we resampled (with replacement) a set of 1,000 simulations from these calibrated models. The final sample included 113 unique parameter sets (shown in Figure 2 in the main text).

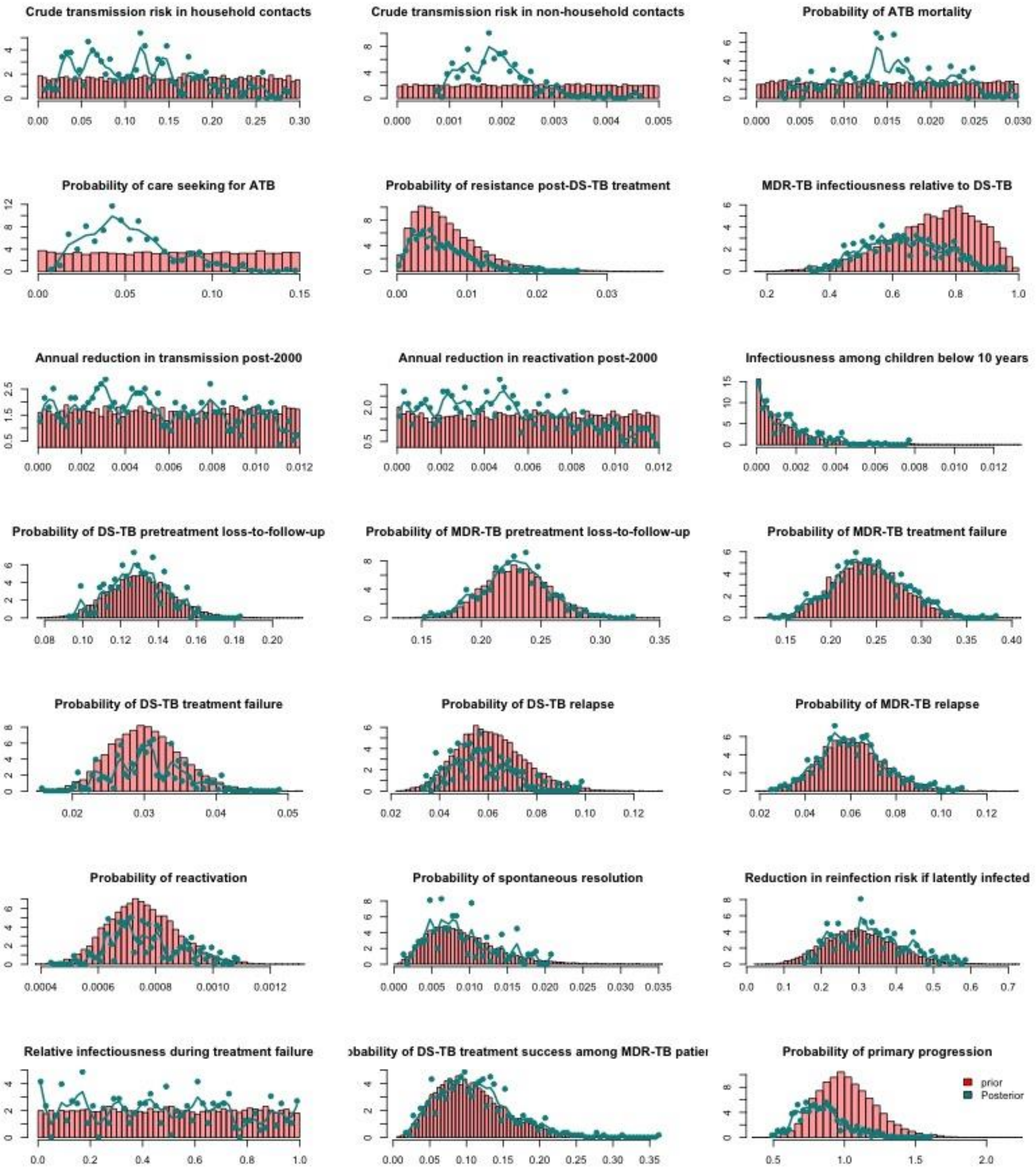

**Figure S5: Sampling distributions of top calibrated simulations.** The red bars represent the prior sampling distribution compared againsed resampled posterior distribution shown in green. Each green dot represents a single calibrated simulation among the top 555 models (accounting for 99.99% of the weighted joint likelihood), and the green line is smoothed spline to characterize the shape of the posterior distribution. The uniform prior distributions were set empirically and the ranges were extended after initiatl experimentation with the model such that the posteriors were not too close to the edge of the prior.

##### 1.5.5 Sampling importance resampling

After the calibration process described above, we identified those highest-ranked simulations for which the sum of their likelihoods comprised 99.99% of the sum of the likelihoods for the full sample; this corresponded to the top 555 simulations. Using the likelihood values as resampling weights, in the next step, we generated a set of 1,000 simulations in year 2022, by randomly sampling (with replacement) available simulations according to weights. The final simulations (sample of 1,000) included 113 unique parameter sets (some parameter sets with low likelihood values were not resampled, and some with high log-likelihood values were resampled multiple times). **Figure 2** in the main text represent the calibration fit among these 1,000 resampled models. Each simulated model was replicated 50 times under 4 different TPT scenarios from 2023 to 2040.

#### 1.6 Experimental scenarios

The main intervention scenario represents a household contact tracing (HHCT) program for people diagnosed with active MDR-TB, which includes screening of household members for active and latent TB and, potentially, treating diagnosed infections. The intervention begins in year 2023 and continues to year 2027. The population is followed to year 2040 to project the mid-term impact on main epidemiological outcomes.

Specifically, we trace a proportion of individuals who are initiating treatment for MDR-TB (those diagnosed with MDR-TB who do not experience pre-treatment loss to follow up) to their households. We exclude household members who are receiving treatment for active DS or MDR-TB at the time of the intervention. All other household members are screened for active TB using a test with imperfect sensitivity, and those diagnosed with active disease receive appropriate treatment (i.e., given the known MDR-TBcontact, all receive DST and an appropriate treatment regimen). Assuming that a combination of clinical evaluation and Xpert is used for detecting active infections, we estimate a 90% sensitivity for new diagnosis.

The remaining household members are screened for latent TB infection (**Figure 1-left**). We assume a sensitivity of 90% for detecting latent infections. Those diagnosed with latent TB are assumed to be treated with a 6-month preventive therapy (PT) regimen. We further assume that children under the age of 5 will receive preventive therapy without the need for a test for latent TB. We compare three different intervention scenarios, with different TPT regimens: 1) a TPT regimen (e.g., isoniazid) with activity only against DS-TB infections, 2) a TPT regimen (e.g., delamanid) with activity against both DS-TB and DR-TB infections, and 3) no TPT/placebo (as a comparator arm, with no effect on TB infections).

To improve interpretability, we simulate only one HHCT event per household (i.e., we do not repeat testing or treatment initiation if other household members are diagnosed with TB after the initial contact investigation event in a given household). Furthermore, to ensure that we simulate the full impact of preventive therapy on TB transmission in households, we track household membership until no household members are taking preventive therapy (e.g., at the planned household dissolution time 10 years after the most recent active TB onset, we check the household members, and if someone is on TPT, we extend the follow-up by one month, and check the status again until no household member is taking TPT).

##### 1.6.1 Impact of preventive therapy:

PT is modeled as having three effects on TB risk: reduction in risk of progression to active disease (among those with early and late latent infections), reduction in risk of relapse (among those recently recovered from TB), and additional protection against new infections/reinfections (for everyone). All effects start immediately when the TPT regimen starts. The protection against infection/reinfection lasts only for the duration of therapy, whereas the effects on progression and relapse persist after TPT ends.

The impact of TPT using isoniazid is restricted to existing and new DS-TB infections, while delamanid can treat or prevent both DS-TB and MDR-TB infections. The efficacy of TPT against progression of pre-existing infections is modeled as all-or-none in a given individual, but due to a combination of imperfect efficacy and incomplete adherence,

TPT is not successful in all recipients. If successful, the impact of TPT on progression to active TB is modeled as 100% reduction in both primary progression and reactivation that persists indefinitely for infections present at the time of TPT initiation (**Figure 1-right**). These individuals will be moved to the recovered state (experiencing no risk of relapse to active disease and retaining lifetime partial immunity toward future reinfections in the recovered state). Similarly, we model the impact of successful TPT against recently recovered TB as 100% reduction in risk of relapse to active TB for the remaining duration of the two-year recently recovered period. Our primary analyses set the probability of success to 70% for progression infection and 70% for relapse of recently-recovered disease; in sensitivity analysis, we vary these two probabilities independently between 20% and 100%]. Finally, we assume that all individuals receiving TPT (regardless of TPT success or failure) will experience a partial reduction in risk of infection/reinfection if exposed to someone with active TB while taking TPT.

For individuals who have active TB disease while they are receiving TPT (due to undetected active TB at the start of TPT, progression from latent to active disease despite TPT, or new infection or reinfection despite TPT), TPT is assumed to have no impact on disease progression, care seeking and diagnosis, or treatment outcomes. TPT will be discontinued when active TB is diagnosed. **Table S4** lists the parameters used to inform the experimental scenarios and their corresponding ranges used in sensitivity analysis.

**Table S4: List of intervention parameters, baseline values and ranges for sensitivity analysis**

| Parameter | Baseline value | Range for sensitivity analysis | Reference |
|---|---|---|---|
| Probability of household contact tracing for MDR-TB patients receiving treatment | 70% | 50%-100% | Assumed |
| ATB screening sensitivity | 90% | 50%-100% | [30] |
| LTB screening sensitivity | 90% | 50%-100% | [31,32] |
| Probability of TPT success in clearing latent disease | 70% | 50%-100% | Assumption |
| Probability of TPT success in clearing recently-recovered disease | 70% | 50%-100% | Assumed equal to efficacy against LTBI progression |
| TPT duration | 6 months | | Based on NCT03568383 |
| TPT protection against reinfection | 70% | 50%-100% | Assumed equal to efficacy against LTBI progression |

#### 1.7 Implementation of model

The simulation model is written in C/C++ and is released under the GPL 2.0 Public License. The source code is available at https://github.com/TB-Modeling/mdrtb. The git repository contains the version controls for previous revisions to this model. Version 2 (saved on Branch TPT-GPL2) was used to produce the results in this manuscript.

#### 2 SUPPLEMENTARY RESULTS TABLES/FIGURES

##### 2.1 Projected demographic and epidemiological outcomes under the baseline scenario with no intervention

**Table S5: Projected demographic and epidemiological outcomes under the baseline scenario with no intervention at year 2022 (baseline) and year 2040 (end of projections).** Values represent the median [interquartile range (IQR)] of simulated outcomes.

| | Year 2022 | Year 2040 |
|---|---|---|
| **Demographic outcomes** | | |
| Population size | 794,295 [794,013 – 794,295] | 899,351 [899,032 – 899,351] |
| Median age of population (years) | 27 [27 – 27] | 33 [33 – 33] |
| Population proportion ≤5 years old | 0.10 [0.10 – 0.11] | 0.85 [ 0.85 – 0.86] |
| Population proportion ≤15 years old | 0.28 [0.28 – 0.29] | 0.23 [0.23 – 0.24] |
| Median age of people in households (years) | 27 [27 – 27] | 28 [28 – 28] |
| Median size of households | 5 [5 – 5] | 5 [5 – 5] |
| **Epidemiological outcomes** | | |
| TB prevalence (per 100k persons) | 280 [267 – 288] | 109 [106 – 144] |
| TB mortality rate (per 100k/year) | 42 [40 – 43] | 17 [15 – 19] |
| TB incidence rate (per 100k/year) | 149 [143 – 164] | 57 [56 – 79] |
| Proportion of TB incidence originating from household transmission | 0.24 [0.24 – 0.32] | 0.23 [0.23 – 0.30] |
| DS prevalence (per 100k persons) | 269 [257 – 276] | 103 [101 – 138] |
| DS mortality rate (per 100k/year) | 40 [38 – 41] | 16 [14 – 18] |
| DS incidence rate (per 100k/year) | 144 [140 – 158] | 55 [54 – 77] |
| Proportion of DS-TB incidence originating from household transmission | 0.25 [0.24 – 0.33] | 0.23 [0.23 – 0.30] |
| Proportion of DS-TB incidence due to primary progression | 0.83 [0.82 – 0.83] | 0.67 [0.65 – 0.70] |
| Proportion of DS-TB incidence due to reactivation | 0.13 [0.12 – 0.13] | 0.28 [0.25 – 0.3] |
| Proportion of DS-TB incidence due to relapse | 0.04 [0.04 – 0.05] | 0.05 [0.04 – 0.05] |
| MDR-TB prevalence (per 100k persons) | 11 [10 – 11] | 5 [5 – 6] |
| MDR-TB mortality rate (per 100k/year) | 2 [2 – 2] | 0.9 [0.68 – 1] |
| MDR-TB incidence rate (per 100k/year) | 5 [4 – 5] | 2 [2 – 2] |
| Proportion of MDR-TB incidence originating from household transmission | 0.24 [0.18 – 0.32] | 0.22 [0.2 – 0.26] |
| Proportion of MDR-TB incidence due to primary progression | 0.79 [0.69 – 0.82] | 0.60 [0.51 – 0.64] |
| Proportion of MDR-TB incidence due to reactivation | 0.1 [0.09 – 0.11] | 0.26 [0.24 – 0.28] |
| Proportion of MDR-TB incidence due to relapse | 0.01 [0.01 – 0.04] | 0.04 [0.04 – 0.05] |
| Proportion of MDR-TB incidence due to newly acquired resistance | 0.08 [0.08 – 0.19] | 0.09 [0.06 – 0.19] |

MDR-TB: multidrug-resistant tuberculosis; DS-TB: drug-susceptible tuberculosis;

#### 2.2 Projected programmatic outcomes of contact investigation for household members of people diagnosed with MDR-TB

**Table S6: Projected programmatic outcomes of contact investigation for household members of people diagnosed with MDR-TB.**

| Cumulative outcomes, 2023 – 2027[1] | Median [interquartile range] |
|---|---|
| Incident cases of MDR-TB | 169 [134 – 178] |
| Incident MDR-TB cases that are notified | 123 [112 – 127] |
| MDR-TB cases whose households complete HHCT | 65 [53 – 68] |
| Number of household members screened | 314 [255 – 327] |
| Number of prevalent active TB cases diagnosed during HHCT | 10 [8 – 11] |
| Prevalent DS-TB cases | 2 [2 – 2] |
| Prevalent MDR-TB cases | 8 [6 – 9] |
| Household contacts starting TPT | 211 [175 – 225] |
| Number of TPT recipients ≤5 years old | 34 [29 – 36] |
| Number of TPT recipients > 5 years old with evidence of TB infection | 176 [146 – 189] |
| Latent DS-TB | 114 [108 – 125] |
| Latent MDR-TB | 110 [86 – 128] |
| Latent DS-TB & MDR-TB | 49[ 47 – 64] |

[1] Cumulative five-year outcomes among simulated populations with an average size of 800,000 people in 2022.

MDR-TB: multidrug-resistant tuberculosis; DS-TB: drug-susceptible tuberculosis; HHCT: household contact tracing; TPT: TB prevention therapy

#### 2.3 Projected impact of TPT scenarios among TPT recipients over short-term and long-term

Using a similar approach to a clinical trial of a TPT intervention, we followed each TPT-eligible contacts for two years (or until the first event of TB incidence or death) and projected the cumulative outcomes in each scenario (**Table S7**). To estimate long-term impact, we further followed TPT-eligible contacts to year 2040 (median of 16 years of follow up).

**Table S7: Projected impact of TPT scenarios among TPT recipients over 2 year and a median 16.17 [IQR 14.58 –17.67] years follow up**

| | | | | Incident cases prevented relative to placebo scenario[1] | | | | |
|---|---|---|---|---|---|---|---|---|
| | DS-TB incidence per TPT recipient | MDR-TB incidence per TPT recipient | Total active TB incidence per TPT recipient | DS-TB | MDR-TB | Total active TB | NNT to avert one MDR-TB incident case | NNT to avert one active (DS or MDR) TB incident case |
| **Events projected among TPT recipients over 2 years** | | | | | | | | |
| Placebo scenario | 0.4% [0 – 0.8%] | 2% [1.3 – 2.9%] | 2.5% [1.6 – 3.5%] | - | - | - | - | - |
| Isoniazid scenario | 0% [0 – 0.4%] | 1.3% [2.9 – 2%] | 1.6% [3.5 – 2.2%] | 100% [59 - 100%] | 3% [-54 - 41%] | 14% [-34 - 46%] | Inf [104 – Inf] | 342 [74 - Inf] |
| Delamanid scenario | 0% [0 – 0.3%] | 2.9% [2 – 1.3%] | 3.5% [2.2 – 1.4%] | 100% [61 - 100%] | 72% [45 - 100%] | 70% [46 - 88%] | 73 [44 – 176] | 60[37-130] |
| **Events projected among TPT recipients over 16 years** | | | | | | | | |
| Placebo scenario | 1.7% [0.9 – 2.6%] | 3.8% [2.6 – 5.4%] | 5.7% [4.1 – 7.7%] | - | - | - | - | - |
| Isoniazid scenario | 1.1% [0.5 – 1.8%] | 2.6% [5.4 – 3.8%] | 4.1% [7.7 – 5%] | 43% [-12 - 76%] | 0% [-49 - 34%] | 11% [-26 - 37%] | Inf [66 – Inf] | 166 [40 - Inf] |
| Delamanid scenario | 1.1% [0.5 – 1.8%] | 5.4% [3.8 – 2.6%] | 7.7% [5 – 3.6%] | 42% [-13 - 75%] | 51% [20 - 72%] | 45% [18 - 64%] | 54 [30 – 183] | 40 [23 - 124] |

[1]Simulations with no incident DS-TB in the Placebo scenario are reported as 100% prevented.

#### 2.4 Projected impact of TPT scenarios at the population level

To estimate the impact of TPT on the broader population, we followed the population to year 2040 and projected the cumulative outcomes in each scenario (**Table S8)**. In contrast to estimates above (Table S7), we did not exclude individuals upon first event of infection with TB and followed everyone to death or year 2040.

**Table S8: Projected impact of TPT scenarios at the population level**

| | | | | Incident cases prevented relative to placebo scenario | | | | |
|---|---|---|---|---|---|---|---|---|
| | DS-TB incident per TPT recipient | MDR-TB incident per TPT recipient | Total active TB incidence per TPT recipient | DS-TB | MDR-TB | Total active TB | NNT to avert one MDR-TB incident case | NNT to avert one active (DS or MDR) TB incident case |
| **Events projected among full population over 18 years** | | | | | | | | |
| No intervention[1] | 68.4 [65.5 – 84.8] | 2.3 [1.9 – 2.4] | 70.4 [67.1 – 87.2] | - | - | - | - | - |
| Placebo scenario | 68.2 [65.5 – 85.4] | 2.2 [1.9 – 2.4] | 70.2 [67.7 – 87.6] | - | - | - | - | - |
| Isoniazid scenario | 68.3 [65.5 – 85.4] | 2.2 [1.9 – 2.4] | 70.2 [67.7 – 88.1] | -0.12% [-2 – 2%] | -0.24% [-2 – 2%] | -0.12% [-2 – 2%] | Inf [24 – Inf] | Inf [1 – Inf] |
| Delamanid scenario | 68.3 [65.5 – 84.7] | 2.2 [1.9 – 2.3] | 70.3 [67.6 – 87.1] | 0.05% [-2 – 2%] | 1.6% [-0.6 – 3.6%] | 0.08% [-2 – 2%] | 27 [11 – Inf] | 18 [1 – Inf] |
| | | | | **Incident cases prevented relative to no intervention** | | | | |
| **Events projected among full population over 18 years** | | | | | | | | |
| No intervention | Same as above | | | - | - | - | - | - |
| Placebo scenario | | | | -0.01% [-2 – 2%] | 2% [-0.37 – 4%] | 0.04% [-2 – 2%] | 22 [10 – Inf] | 42 [1 – Inf] |
| Isoniazid scenario | | | | 0.07% [-2 – 2%] | 2% [-0.56 – 4%] | 0.09% [-2 – 2%] | 22 [10 – Inf] | 14 [1 – Inf] |
| Delamanid scenario | | | | 0.07% [-0.31 – 0.42%] | 3.6% [2 – 5.2%] | 0.19% [-0.19 – 0.52%] | 12 [8 – 22] | 7 [3 – Inf] |

[1] Relative to the number of TPT recipients in the Delamanid scenario

#### 2.5 Projected TB outcomes at the population level by year 2040 under alternative scenarios

The projected MDR-TB incidence in 2040 decreased from 2.1 [IQR 1.93 – 2.38] cases per 100,000 persons with no household intervention to 2.04 [IQR 1.88 – 2.31] when household contacts received TB screening and delamanid TPT from 2023 – 2027, a 3% [IQR -1 – 6%] reduction. (This is smaller than the 8% [IQR 4 – 12%] reduction in cumulative incidence reported in the main Results, because most of the averted MDR-TB cases were averted in earlier years, and incidence had partly returned to baseline by 2040, 13 year after the household intervention ended.)

**Table S9** represents projected total events per TPT recipient by year 2040 at the population-level. Values represent the Median [interquartile range (IQR)] of simulated outcomes.

**Table S9: Projected total TB outcomes at the population level, 2023 – 2040**

| | Number of events per one TPT recipient | | | |
|---|---|---|---|---|
| **Projected Outcomes** | **No intervention[1]** | **Placebo Scenario** | **Isoniazid Scenario** | **Delamanid Scenario** |
| Total TB incidence | 70.43 [67.77 – 87.17] | 70.22 [67.72 – 87.63] | 70.24 [67.74 – 88.1] | 70.27 [67.64 – 87.06] |
| DS incidence | 68.42 [65.53 – 84.82] | 68.2 [65.48 – 85.37] | 68.31 [65.48 – 85.39] | 68.33 [65.48 – 84.74] |
| DS incidence due to primary progression | 52.46 [50.12 – 65.63] | 52.3 [50.13 – 65.19] | 52.29 [50.1 – 65.56] | 52.42 [50.07 – 65.62] |
| DS incidence due to reactivation | 12.67 [12.24 – 13.85] | 12.66 [12.22 – 13.8] | 12.66 [12.24 – 13.89] | 12.66 [12.23 – 13.87] |
| DS incidence due to relapse | 3.18 [3.07 – 3.61] | 3.18 [3.06 – 3.61] | 3.18 [3.06 – 3.59] | 3.18 [3.06 – 3.59] |
| MDR incidence | 2.29 [1.95 – 2.44] | 2.24 [1.89 – 2.37] | 2.25 [1.91 – 2.37] | 2.21 [1.88 – 2.34] |
| MDR incidence due to primary progression | 1.64 [1.15 – 1.76] | 1.6 [1.12 – 1.7] | 1.61 [1.11 – 1.7] | 1.56 [1.09 – 1.67] |
| MDR incidence due to reactivation | 0.395 [0.307 – 0.418] | 0.392 [0.305 – 0.418] | 0.394 [0.304 – 0.417] | 0.392 [0.304 – 0.414] |
| MDR incidence due to relapse | 0.071 [0.066 – 0.12] | 0.07 [0.066 – 0.118] | 0.07 [0.066 – 0.118] | 0.069 [0.065 – 0.116] |
| MDR incidence due to resistance | 0.238 [0.16 – 0.382] | 0.235 [0.16 – 0.381] | 0.239 [0.161 – 0.381] | 0.237 [0.161 – 0.382] |
| DS Mortality | 18.89 [18.11 – 20.35] | 18.93 [18.02 – 20.28] | 18.95 [18.02 – 20.25] | 18.9 [18.1 – 20.34] |
| MDR Mortality | 0.993 [0.907 – 1.04] | 0.967 [0.885 – 1.01] | 0.97 [0.887 – 1.01] | 0.953 [0.877 – 0.996] |
| DS incidence originating from household transmission | 16.31 [15.61 – 24.93] | 16.27 [15.63 – 24.98] | 16.25 [15.62 – 24.87] | 16.3 [15.61 – 24.81] |
| MDR incidence originating from household transmission | 0.526 [0.454 – 0.616] | 0.512 [0.443 – 0.594] | 0.512 [0.444 – 0.599] | 0.489 [0.427 – 0.561] |
| DS/MDR incidence originating from household transmission | 16.69 [16.11 – 25.58] | 16.67 [16.1 – 25.67] | 16.64 [16.11 – 25.59] | 16.67 [16.07 – 25.52] |

[1] Relative to the number of TPT recipients in the Delamanid scenario

MDR-TB: multidrug-resistant tuberculosis; DS-TB: drug-susceptible tuberculosis; TPT: TB prevention therapy

#### 3 SENSITIVITY ANALYSIS

##### 3.1 Sensitivity to core model parameters

For each of the 21 key simulation parameters (**Table 1**), we calculated partial rank correlation coefficients (PRCCs) to determine the association between that parameter and projected reductions in cumulative MDR-TB incidence via delamanid TPT (**Figure S6**). Those parameters most strongly correlated with this primary outcome (i.e., with absolute value of PRCC>0.15) were selected for one-way sensitivity analysis, in which we compared the subset of simulations with that parameter in its highest quartile to the subset of simulations with that parameter in its lowest quantile (**Figure S7**).

The parameters which were most strongly correlated with the primary outcome included the crude TB transmission risk among household contacts (per month), reduction in reinfection risk if latently infected, probability of ATB mortality, MDR-TB infectiousness relative to DS-TB, and probability of primary progression.

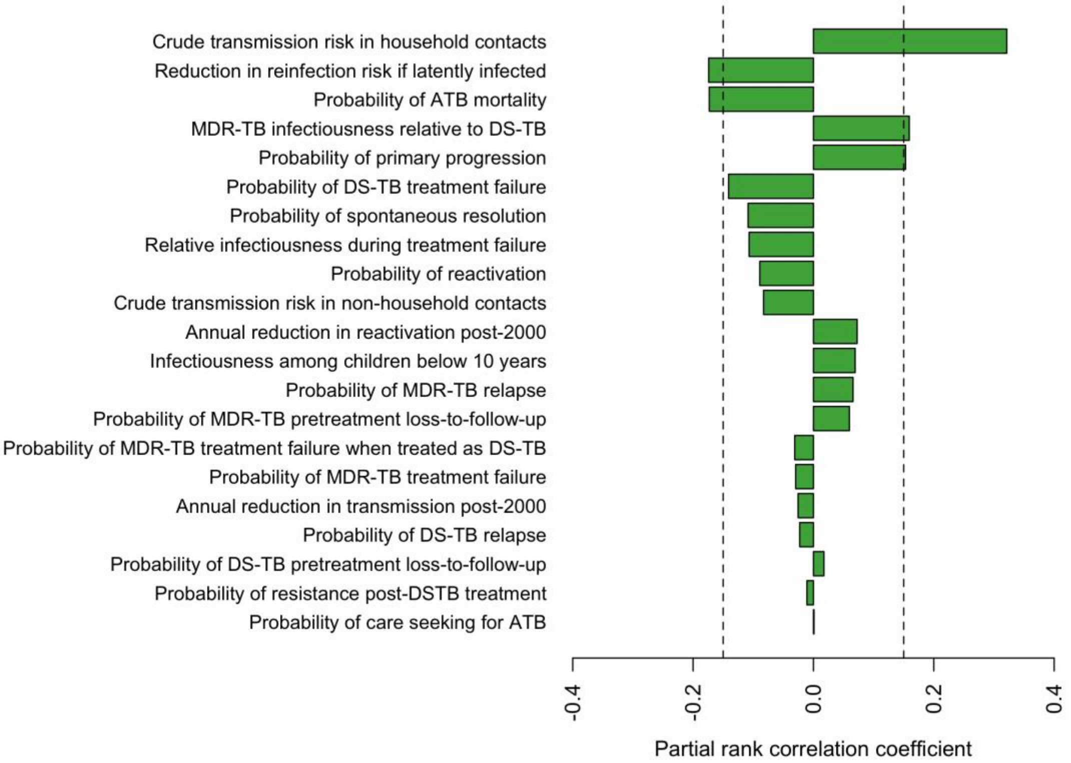

**Figure S6: Partial rank correlation coefficients (PRCCs) for model parameters.** For each model parameter sampled during the calibration processes, we calculated PRCCs to determine the association between that parameter and projected reductions in cumulative MDR-TB incidence with household contact investigation and delamanid TPT. Dashed vertical lines mark the threshold of ±0.15 that we used to identify parameters most strongly associated with the primary outcome.

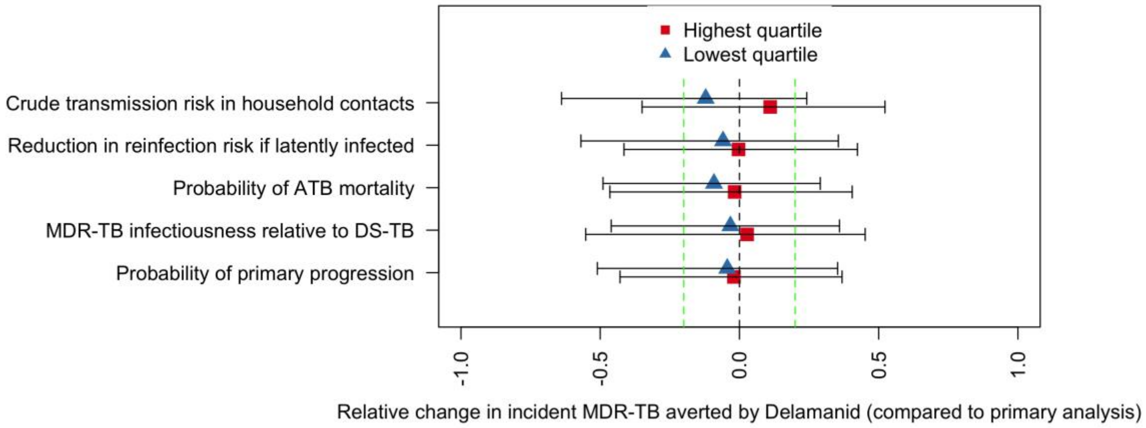

**Figure S7: Sensitivity of the impact of Delamanid impact on MDR-TB incidence to extreme value of core model parameters.** For those parameters most strongly associated with the primary outcome (absolute value of PRCC >0.2), the subset of simulations with that parameter in the highest quartile is compared against simulations with that parameter in the lowest quartile. The primary outcome of interest is the relative change in the impact of Delamanid on cumulative MDR-TB incidence (e.g., 0.5 represents a 50% increase in the the difference in cumulative incident MDR-TB in the population between 2022 and 2040, comparing household screening + TPT to no household intervention). Shapes show median values and whiskers mark the 75% uncertainty ranges. Dashed vertical lines in green mark a threshold of +/-20% to identify epidemiologically significant differences.

##### 3.2 Sensitivity analysis to household intervention parameters

To assess the sensitivity of projected outcomes under alternative TPT configurations, we simulated the delamanid arm across 1000 primary simulation models by randomly sampling each of the six TPT parameters from corresponding prior distributions (**Table S4**). The procedure was repeated for 100 random replications. We compared the sensitivity of primary outcomes (projected cumulative MDR-TB incidence among TPT recipients and at the population level by year 2040) between the subsets of simulations that contained the highest versus lowest quartile of each TPT parameter.

The projected MDR-TB incidence among TPT recipients was most sensitive to the success of TPT in clearing latent disease that could progress to active infections (**Figure S8**). Reductions in ATB screening sensitivity were also associated with an increase in MDR-TB incidence among TPT recipients, because household contacts with undiagnosed active disease experienced no benefit from TPT and continued to transmit TB until successful diagnosis and treatment with an appropriate treatment regimen. At a population level, the probability of household contact tracing and LTB screening sensitivity had the largest impact on projected MDR-TB incidence under the Delamanid arm (**Figure S9**).

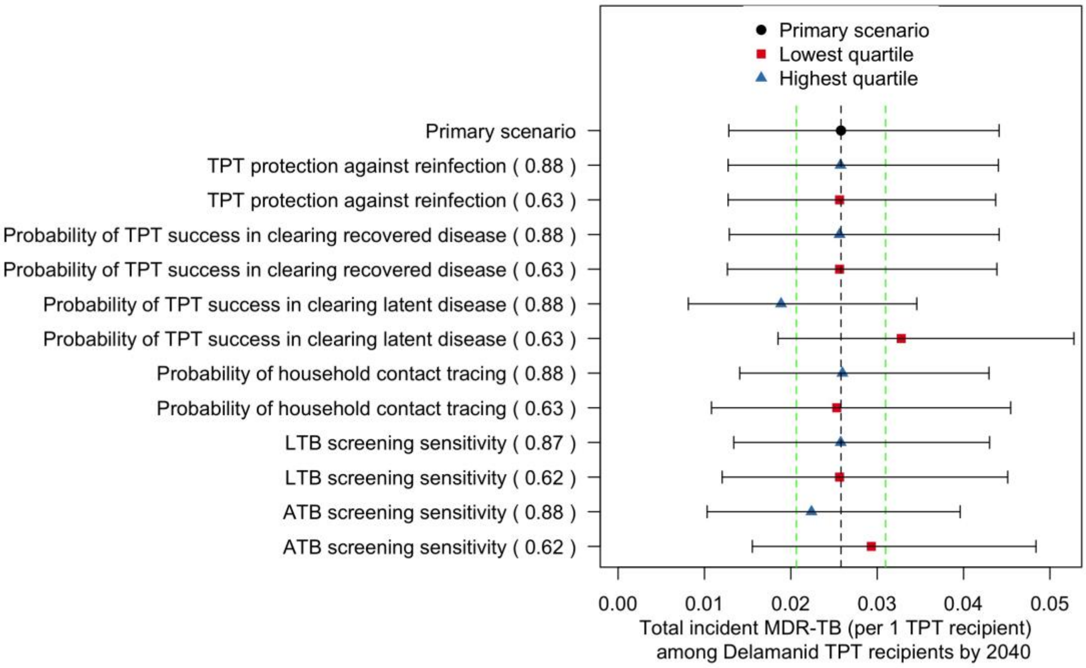

**Figure S8: Sensitivity of impact of Delamanid TPT among recipients to alternative TPT configurations.** The X-axis presents the cumulative MDR-TB incidence (per 1 TPT recipient) among delamanid TPT recipients by year 2040. For each TPT parameter, we compare simulations with that parameter in the highest quartile (marked via blue triangle) against simulations with that parameter in the lowest quartile (marked via red squares). Primary scenario represents simulations using the original TPT parameter values (mark via black circle). Circle, Trianges and squares show median values, and whiskers mark the 75% uncertainty ranges. Dashed vertical lines in green mark a threshold of +/-20% of median output in the primary scenario.

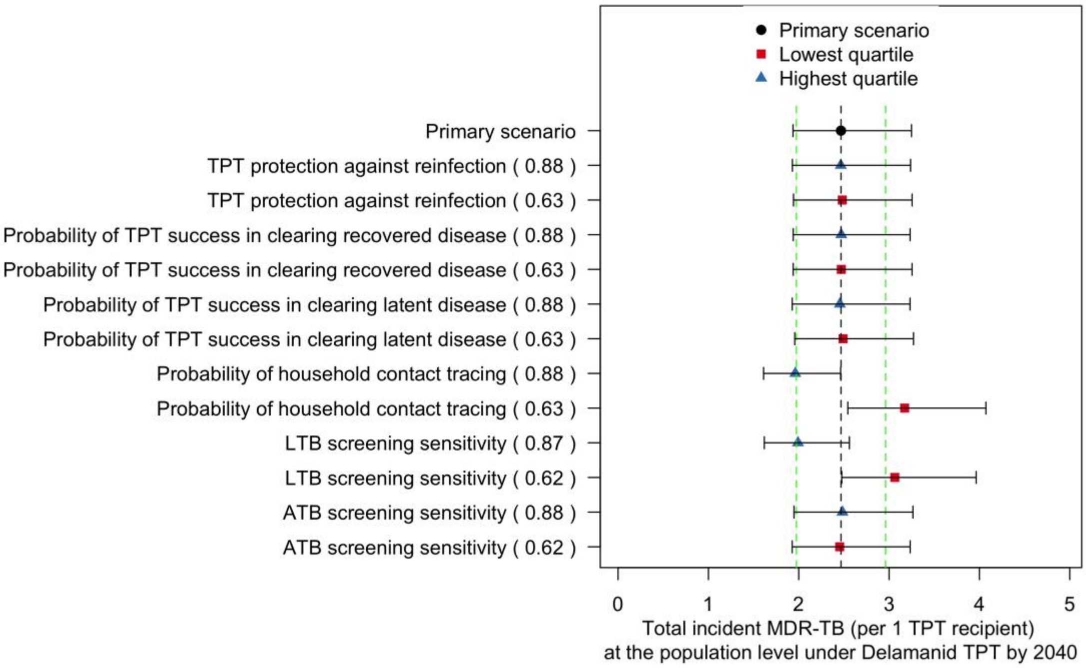

**Figure S9: Sensitivity of impact of Delamanid TPT at the population level to alternative TPT configurations**. The X-axis presents the cumulative MDR-TB incidence (per 1 TPT recipient) at the population level under Delamanid TPT by year 2040. For each TPT parameter, we compare simulations with that parameter in the highest quartile (marked via blue triangle) against simulations with that parameter in the lowest quartile (marked via red squares). Primary scenario represents simulations using the original TPT parameter values (mark via black circle). Circle, Trianges and squares show median values, and whiskers mark the 75% uncertainty ranges. Dashed vertical lines in green mark a threshold of +/-20% of median output in the primary scenario.

**References**

1. United Nations DoEaSA, Population Division;. World Population Prospects 2019, Online Edition. Rev. 1. 2019. https://population.un.org/wpp/Download/Standard/Population/2021).

2. Desai S, Vanneman R. India human development survey-ii (ihds-ii), 2011-12: Inter-university Consortium for Political and Social Research Ann Arbor, MI; 2015.

3. Prem K, Cook AR, Jit M. Projecting social contact matrices in 152 countries using contact surveys and demographic data. *PLoS computational biology* 2017; **13**(9): e1005697.

4. Mossong J, Hens N, Jit M, et al. Social contacts and mixing patterns relevant to the spread of infectious diseases. *PLoS medicine* 2008; **5**(3): e74.

5. Vynnycky E, Fine P. The natural history of tuberculosis: the implications of age-dependent risks of disease and the role of reinfection. *Epidemiology & Infection* 1997; **119**(2): 183-201.

6. Kasaie P, Andrews JR, Kelton WD, Dowdy DW. Timing of tuberculosis transmission and the impact of household contact tracing. An agent-based simulation model. *Am J Respir Crit Care Med* 2014; **189**(7): 845-52.

7. World Health Organization (WHO). Diagnosis, notification and treatment of rifampicin-resistant TB. Geneva, 2021.

8. Starke JR. Transmission of Mycobacterium tuberculosis to and from children and adolescents. Seminars in Pediatric Infectious Diseases; 2001: Elsevier; 2001. p. 115-23.

9. Subbaraman R, Nathavitharana RR, Satyanarayana S, et al. The tuberculosis cascade of care in India's public sector: a systematic review and meta-analysis. *PLoS medicine* 2016; **13**(10): e1002149.

10. MacPherson P, Houben RM, Glynn JR, Corbett EL, Kranzer K. Pre-treatment loss to follow-up in tuberculosis patients in low-and lower-middle-income countries and high-burden countries: a systematic review and meta-analysis. *Bulletin of the World Health Organization* 2013; **92**: 126-38.

11. Fox GJ, Barry SE, Britton WJ, Marks GB. Contact investigation for tuberculosis: a systematic review and meta-analysis. *European Respiratory Journal* 2013; **41**(1): 140-56.

12. Horsburgh Jr CR. Priorities for the treatment of latent tuberculosis infection in the United States. *New England Journal of Medicine* 2004; **350**(20): 2060-7.

13. Dale KD, Trauer JM, Dodd PJ, Houben RM, Denholm JT. Estimating long-term tuberculosis reactivation rates in Australian migrants. *Clinical Infectious Diseases* 2020; **70**(10): 2111-8.

14. Tiemersma EW, van der Werf MJ, Borgdorff MW, Williams BG, Nagelkerke NJ. Natural history of tuberculosis: duration and fatality of untreated pulmonary tuberculosis in HIV negative patients: a systematic review. *PloS one* 2011; **6**(4): e17601.

15. Ragonnet R, Flegg JA, Brilleman SL, et al. Revisiting the natural history of pulmonary tuberculosis: a bayesian estimation of natural recovery and mortality rates. *BioRxiv* 2019: 729426.

16. Andrews JR, Noubary F, Walensky RP, Cerda R, Losina E, Horsburgh CR. Risk of progression to active tuberculosis following reinfection with Mycobacterium tuberculosis. *Clinical infectious diseases* 2012; **54**(6): 784-91.

17. Marx FM, Dunbar R, Enarson DA, et al. The temporal dynamics of relapse and reinfection tuberculosis after successful treatment: a retrospective cohort study. *Clinical infectious diseases* 2014; **58**(12): 1676-83.

18. Ahuja SD, Ashkin D, Avendano M, et al. Multidrug resistant pulmonary tuberculosis treatment regimens and patient outcomes: an individual patient data meta-analysis of 9,153 patients. *PLoS medicine* 2012; **9**(8): e1001300.

19. Geneva: World Health Organization. GLOBAL TUBERCULOSIS REPORT 2019, 2019.

20. O'Donnell MR, Padayatchi N, Kvasnovsky C, Werner L, Master I, Horsburgh Jr CR. Treatment outcomes for extensively drug-resistant tuberculosis and HIV co-infection. *Emerging infectious diseases* 2013; **19**(3): 416.

21. Salvatore PP, Proaño A, Kendall EA, Gilman RH, Dowdy DW. Linking individual natural history to population outcomes in tuberculosis. *The Journal of infectious diseases* 2018; **217**(1): 112-21.

22. Grandjean L, Gilman RH, Martin L, et al. Transmission of multidrug-resistant and drug-susceptible tuberculosis within households: a prospective cohort study. *PLoS medicine* 2015; **12**(6): e1001843.

23. Borrell S, Gagneux S. Infectiousness, reproductive fitness and evolution of drug-resistant Mycobacterium tuberculosis [State of the art]. *The International Journal of Tuberculosis and Lung Disease* 2009; **13**(12): 1456-66.

24. Menzies D, Benedetti A, Paydar A, et al. Effect of duration and intermittency of rifampin on tuberculosis treatment outcomes: a systematic review and meta-analysis. *PLoS medicine* 2009; **6**(9): e1000146.

25. Menzies D, Benedetti A, Paydar A, et al. Standardized treatment of active tuberculosis in patients with previous treatment and/or with mono-resistance to isoniazid: a systematic review and meta-analysis. *PLoS medicine* 2009; **6**(9): e1000150.

26. World Health Organization (WHO). Global Tuberculosis Report. Geneva; 2021.

27. Houben RM, Dodd PJ. The global burden of latent tuberculosis infection: a re-estimation using mathematical modelling. *PLoS medicine* 2016; **13**(10): e1002152.

28. Martinez L, Shen Y, Mupere E, Kizza A, Hill PC, Whalen CC. Transmission of Mycobacterium tuberculosis in households and the community: a systematic review and meta-analysis. *American journal of epidemiology* 2017; **185**(12): 1327-39.

29. World Health Organization (WHO). National TB Prevalence Survey in India 2019-2021. Geneva; 2022.

30. Dorman SE, Schumacher SG, Alland D, et al. Xpert MTB/RIF Ultra for detection of Mycobacterium tuberculosis and rifampicin resistance: a prospective multicentre diagnostic accuracy study. *The Lancet infectious diseases* 2018; **18**(1): 76-84.

31. Oh CE, Ortiz-Brizuela E, Bastos ML, Menzies D. Comparing the diagnostic performance of QuantiFERON-TB gold plus to other tests of latent tuberculosis infection: a systematic review and meta-analysis. *Clinical Infectious Diseases* 2021; **73**(5): e1116-e25.

32. Laurenti P, Raponi M, De Waure C, Marino M, Ricciardi W, Damiani G. Performance of interferon-γ release assays in the diagnosis of confirmed active tuberculosis in immunocompetent children: a new systematic review and meta-analysis. *BMC infectious diseases* 2016; **16**(1): 1-11.
